# Lead-in therapy targeting PD1 and/or LAG3 imposes distinct immune phenotypes in first-line treatment of metastatic melanoma

**DOI:** 10.1101/2025.11.04.25339499

**Authors:** Lilit Karapetyan, Anthony R. Cillo, Shuaichao Wang, Arivarasan Karunamurthy, Ryan C. Massa, Anjali Rohatgi, Christopher Deitrick, Yana G. Najjar, Diwakar Davar, Jason J. Luke, Cindy Sander, Sheryl R. Kunning, Amy Rose, Sarah Bradley, Elizabeth Rush, Marion Joy, Riyue Bao, Hong Wang, Dario A.A. Vignali, Tullia C. Bruno, John M. Kirkwood

## Abstract

Simultaneous blockade of lymphocyte activation gene-3 (LAG-3) and programmed-death-1 (PD-1) pathways enhance anti-tumor activity in patients with melanoma. It is not known if this immunotherapy induces unique immune modulation or how this may relate to clinical outcomes, compared to the single modalities. We conducted a randomized three-arm phase 2 trial (NCT03743766) in advanced melanoma comparing lead-in treatment with one cycle of relatlimab-(n=14), nivolumab-(n=15), and nivolumab-relatlimab (n=14) as first-line therapy, followed by combination nivolumab-relatlimab, to assess the impact of the lead-in therapy on immune populations, related to objective response rate, progression-free survival (PFS) and major pathologic response on biopsy (MPRbx). Unexpectedly, diminished efficacy was observed when nivolumab or relatlimab was given as a lead-in monotherapy, despite receipt of the combination subsequent to week 4. MPRbx at week 4 was significantly higher with nivolumab- and combination *vs.* relatlimab-lead-in, mechanistically correlated with higher IFN-g and TCR signaling in CD8^+^ T cells, and was associated with superior PFS. Relatlimab lead-in was characterized by clustering of CD8^+^ T and FOXP3^+^ T regulatory cells in the tumor microenvironment, enrichment in inhibitory receptor pathways and associated with worse PFS. Combination therapy increased the fraction of CD8^+^T_EM_ cells in association with response to therapy while patients with progressive disease demonstrated decrease in CD14^+^CD16^-^HLA-DR^low^CD33^dim^ monocytic populations. The analyses of component agents afforded by this novel lead-in trial has identified differential clinical and immunological modulation of anti-PD1 and anti-LAG3 in comparison to the combination therapy.

**Highlights:**

- Lead-in monotherapy with nivolumab or relatlimab reduces efficacy to the combo
- MPRbx serves as an early surrogate marker of response and long-term PFS
- Relatlimab lead-in led to CD8^+^ T cell:Treg clustering in the TME and poor PFS
- Combo therapy non-responders had decreased CD14^+^CD16^-^HLA-DR^low^CD33^dim^ monocytes

## Introduction

Treatment with immune checkpoint blockade significantly improves outcomes of patients with advanced melanoma. Long-term follow-up results of CheckMate-067 demonstrate durable benefit of combination anti-programmed-death-1 (PD-1) and anti-cytotoxic T lymphocyte antigen-4 (CTLA-4) therapy in the first-line setting for advanced melanoma.^1^ While efficacy of this regimen is impressive, grade 3 and higher treatment-related adverse events developed in 59% of patients, highlighting the need for safer approaches.^1,2^ Lymphocyte activation gene-3 (LAG-3) is a co-inhibitory receptor expressed on activated CD4^+^ and CD8^+^ T cells. Co-expression of LAG-3 with PD-1 in melanoma TIL in the B16-F10 model, and correlation of blood LAG-3^+^CD8^+^ immune cells with decreased immune checkpoint blockade (ICB) efficacy in melanoma patients led to the hypothesis that co-inhibition of these pathways may improve responses over single-agent anti-PD-1 therapy.^3–5^ Relatlimab is a fully human IgG4 antibody that binds to human LAG-3. Efficacy of nivolumab-relatlimab was reported in a global phase 3 trial for advanced melanoma (RELATIVITY-047), confirming superiority of this combination over nivolumab alone. ^6–8^ In contrast nivolumab-relatlimab subsequent to disease progression on anti-PD-1-based therapy demonstrated limited activity with ORR 9-12% and median PFS 2-3 months in the RELATIVITY-020 trial.^9^ The impact of monotherapy with anti-LAG-3 compared to anti-PD1 or the combination on outcome has not been previously evaluated.

Furthermore, the impact of lead-in monotherapy on combinatorial immunotherapy has not been assessed. Single-agent relatlimab development in the initial phase 1 trials showed limited efficacy in heavily treated patients, but its impact as first-line therapy in advanced melanoma has not been evaluated. Immunological changes driven by LAG-3 and PD-1 directed therapies, and whether they are unique or overlapping has also not been elucidated.

This phase 2 trial evaluated the contributions of nivolumab-, relatlimab-, and nivolumab-relatlimab given as lead-in therapy, followed by the combination in first-line treatment of patients with advanced melanoma (ClinicalTrials.gov: NCT03743766). Here we report efficacy and the differential impact of lead-in therapy for 4 weeks with the single agents or the combination upon long-term outcomes, as well as the first assessment of the effects of relatlimab as a single agent in treatment-naïve metastatic melanoma. When this study was conceptualized, the efficacy of nivolumab-relatlimab was not known and the superiority of the combination versus nivolumab monotherapy had not been reported. Prior to this trial, there were no data regarding the antitumor efficacy of relatlimab in the first-line setting of advanced melanoma. There has been significant interest in the unique immunological impact of LAG-3, and how this might compare with anti-PD-1. Therefore, this study was designed to dissect the immunological impact of anti-LAG-3 and anti-PD-1 versus the combination using a lead-in trial design to interrogate the separate contributions of those agents. To assess distinct immunological changes in the tumor microenvironment and peripheral blood with each lead-in arm, serial correlative studies were performed, including analysis of major pathologic response in week 4 biopsies after lead-in therapy, multiplex immunofluorescence analysis and spatial co-clustering assessment of immune cells in the tumor microenvironment at week 4, and multicolor flow cytometry on peripheral blood mononuclear cells (PBMCs) obtained at baseline and weeks 4 and 16.

## Results

### Patient enrollment and characteristics

Sixty-one patients were screened between 03/2019 and 07/2023 for this phase 2 open-label randomized single-center clinical trial (**Fig. S1A, B**). Forty-three patients were eligible and enrolled. Patients were randomized 14, 15, and 14 to receive relatlimab-, nivolumab- or combination-lead-in therapy, respectively. As of database lock on June 30^th^, 2025, 100% (43/43) had discontinued treatment with the most common reason being disease progression in 42% (18/43). The demographic and clinical characteristics of the patients are shown in **Table 1**. Minor imbalances of disease characteristics amongst the 3 lead-in arms were balanced across the 3 arms, reducing the likelihood that high-risk features influenced our analysis of overall efficacy.

**Table 1.** Baseline demographic and disease characteristics.

| Characteristic * | Total | Relatlimab | Nivolumab | Combination |
| --- | --- | --- | --- | --- |
| Number of Patients (%) | 43 (100%) | 14 (32.6%) | 15 (34.9%) | 14 (32.6%) |
| Age, median, years | 67 | 70 | 69 | 60 |
| Gender number (%) |  |  |  |  |
| Male | 28 (65.1%) | 10 (71.4%) | 8 (53.3%) | 10 (71.4%) |
| Female | 15 (34.9%) | 4 (28.6%) | 7 (46.7%) | 4 (28.6%) |
| ECOG Performance Status |  |  |  |  |
| 0 | 34 (79.1%) | 13 (92.9%) | 10 (66.7%) | 11 (78.6%) |
| 1 | 9 (20.9%) | 1 (7.1%) | 5 (33.3%) | 3 (21.4%) |
| LDH |  |  |  |  |
| < ULN | 6 (14%) | 1 (7.1%) | 3 (20%) | 2 (14.3%) |
| > ULN | 37 (86%) | 13 (92.9%) | 12 (80%) | 12 (85.7%) |
| *BRAF* mutation status |  |  |  |  |
| *BRAF* v600 mutant | 18 (47.4%) | 7 (63.6%) | 5 (35.7%) | 6 (46.2%) |
| *BRAF* v600 wild | 20 (52.6%) | 4 (36.4%) | 9 (64.3%) | 7 (53.8%) |
| Stage at enrollment |  |  |  |  |
| Stage III Advanced | 8 (18.6%) | 2 (14.3%) | 3 (20%) | 3 (21.4%) |
| Stage IV Advanced | 35 (81.4%) | 12 (85.7%) | 12 (80%) | 11 (78.6%) |
| Metastasis stage |  |  |  |  |
| M0 | 6 (14%) | 2 (14%) | 2 (13%) | 2 (14%) |
| M1a | 10 (23%) | 3 (21%) | 4 (27%) | 3 (21%) |
| M1b | 8 (19%) | 4 (29%) | 4 (27%) | 0 (0%) |
| M1c | 17 (40%) | 4 (29%) | 4 (27%) | 9 (64%) |
| M1d | 2 (4.7%) | 1 (7.1%) | 1 (6.7%) | 0 (0%) |
| Liver Metastasis |  |  |  |  |
| Yes | 14 (33%) | 3 (21%) | 4 (27%) | 7 (50%) |
| No | 29 (67%) | 11 (79%) | 11 (73%) | 7 (50%) |

\*For parameters for which data are missing, the numbers of patients with data are indicated for each regimen and are used as the denominators for calculating percentages

### Clinical Outcomes

The response rate at week 4 per Response Evaluation Criteria in Solid Tumors (RECIST) version 1.1 at week 4 was 9.5% among 42 evaluable patients (one patient missed week-4 scan) (**Fig. 1A**). Partial responses at week 4 were 7.7% (n=1), 20% (n=3), and 0% (0) on relatlimab, nivolumab, and combination lead-in arms, respectively. Intention-to-treat analysis including all randomized patients showed best overall response (BOR) to be complete response (CR)=2% (n=1), PR= 47% (n=20), SD=21% (n=9) and PD=30% (n=13). Best overall response rate (BORR) was 21% (n=3; 90% CI, 6.1%-46.6%), 60% (n=9; 90% CI, 36.0%-80.9%), and 64% (n=9; 90%CI, 39.0%-84.7%) in relatlimab, nivolumab-, and combination lead-in arms, respectively (**Fig. 1B**). After the lead-in therapy, two patients died: one due to nivolumab-related myocarditis, and the other due to disease progression while receiving lead-in relatlimab therapy. Neither of these patients received combination therapy. Among 41 patients who received at least one cycle of subsequent combination therapy, best overall response was partial response (CR/PR) =49% (20), stable disease (SD)=22% (9), and progressive disease (PD)=29% (12). Per-protocol nivolumab-relatlimab efficacy analysis was also conducted using week 4 scans as baseline and week 16 after 3 cycles of combination therapy. Among 37 evaluable patients, the analysis revealed an ORR of 40.5% (n=15; 90% CI, 26.9%-55.4%), with SD of 38% (n=14) and PD of 22% (n=8). Median time from randomization to first response was 16 weeks for the whole cohort and did not vary among lead-in arms. Median duration of response has not been reached [NR] (95% CI 22.4 to NR months) among all patients and was NR, 37.5 months, and NR in relatlimab-, nivolumab-, and combo-arms, respectively (**Fig. 1C, D**). With median follow-up 49.1 months (95% CI 42.6 to 60.0 months), there were 74% (n=32) events (death or progression) for PFS and 47% (n=20) for OS (deaths). Median PFS for the whole cohort was 6.5 months (95% CI, 2.2 to 21.7 months) and 1.9 months (95% CI, 0.9 to NR months), 4.7 months (95%CI 1-NR months), and 14.7 months (95% CI, 9.1-NR months) in relatlimab-, nivolumab-, and nivolumab-relatlimab lead-in arms, respectively **(Fig. 1E, S2A)**. Lead-in relatlimab *vs.* nivolumab-relatlimab was associated with significantly worse PFS (HR=2.64, 95% CI, 1.03 to 6.75, p=0.043), and lead-in nivolumab *vs.* nivolumab-relatlimab demonstrated a strong trend towards worse PFS (HR=2.29, 95% CI, 0.91 to 5.80, p=0.079). Adjusting for *BRAFv600* mutation status, lead-in relatlimab *vs.* nivolumab-relatlimab continued to be associated with worse PFS (HR=3.03, 95% CI, 1.06 to 8.77, p=0.041) **(Fig. S2B)**. The 12-, 24- and 48-month-PFS-rates were 29%, 14%, 14% for relatlimab-, 40%, 20%, 10% for nivolumab, and 64%, 50%, 50% for combination-lead-in arms, respectively **(Fig. 1F)**. Median OS was NR (95% CI, 21.9 to NR months) for the whole cohort and 23.6 months (95% CI, 12.1 to NR months), 41.1 months (95% CI, 19.0 to NR months), and NR (27.7-NR months) for relatlimab, nivolumab, and combination lead-in arms, respectively (**Fig. S2C, D**). In summary, while all patients were planned to receive combination therapy after one cycle of 4 weeks of nivolumab or relatlimab monotherapy, this study has shown that single agent lead-in therapy detrimentally impacts therapeutic efficacy of the combination therapy.

**Fig. 1:**
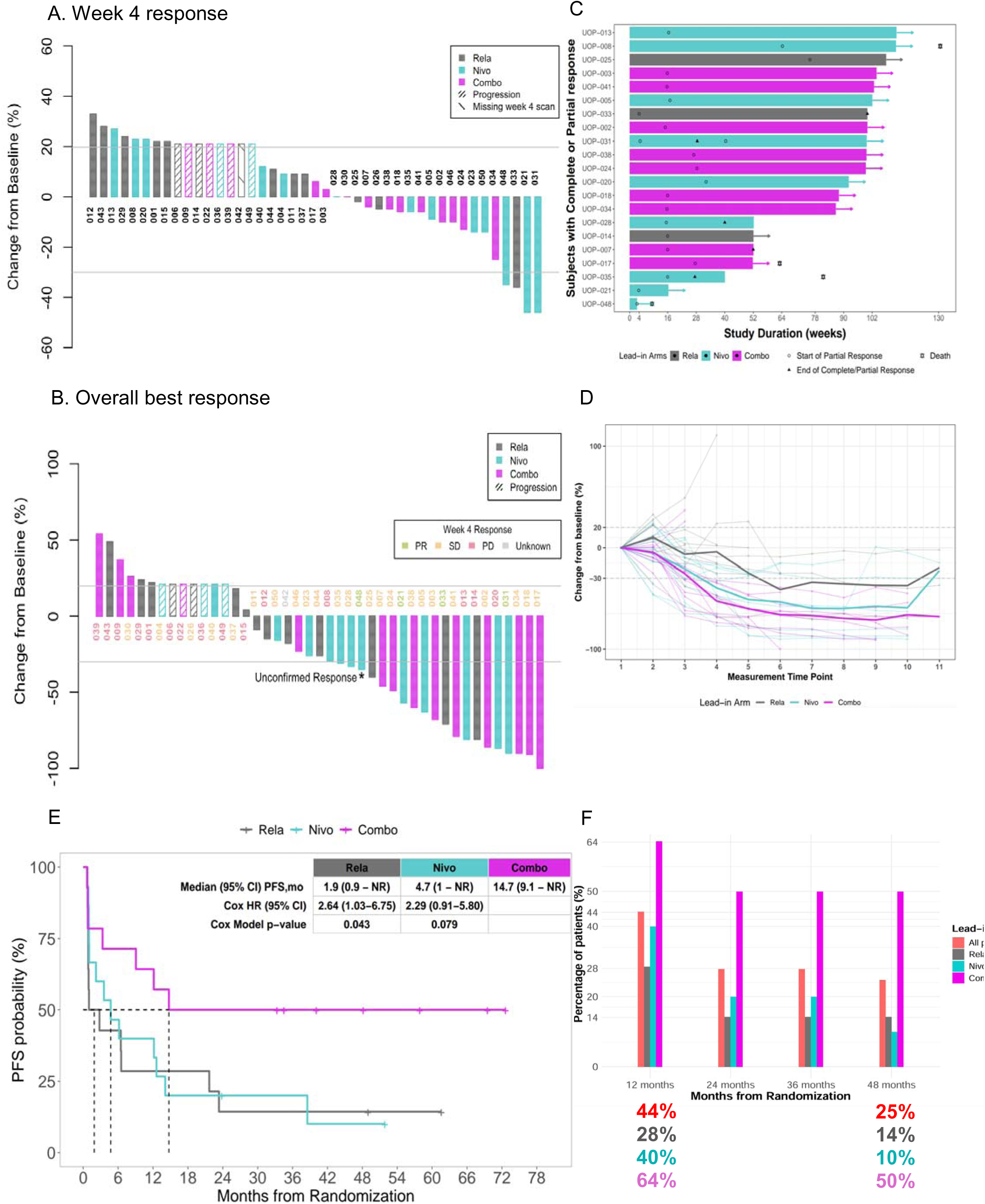
Clinical outcomes of nivolumab, relatlimab, combination lead-in therapy followed by combination therapy. **(A)** Waterfall plot showing change in the sum of target lesion diameters at week 4. Patients with progressive disease due to new lesions, non-target lesions, or early clinical progression are depicted (21% or greater increase) as progression. (**B)** Waterfall plot showing the change in sum of target lesion diameters during follow up in all enrolled patients. (**C)** Swimmer plot depicting individual response, time to development of response, and clinical outcome at data cutoff; **D)** Spider plot showing changes in sum of target lesion diameters throughout treatment. (**E)** PFS according to lead-in arm. **(F)** 12-, 24-, 36-and 48-month-PFS rate among all patients and each lead-in arm. Abbreviations: nivolumab (nivo), relatlimab (rela), and combination (combo).

### Adverse events

Adverse events occurred in ≥10% of overall study patients and were attributed to therapy are listed in **Supplementary table 1, data file S1**. Treatment-related adverse events leading to therapy discontinuation were recorded in 20.9% (n=9) of cases, including colitis (n=2), transaminitis (n=3), bullous pemphigoid (n=1), elevated troponin (n=1), encephalitis (n=1), and one death due to lead-in nivolumab induced myocarditis. Adrenal insufficiency was observed in 16% (n=7) of cases. While the overall safety profile of combination therapy was consistent with the data reported from the pivotal phase III study (RELATIVITY-047),^6,8^ ≥G3 and higher TRAEs were ∼40% among patients in the relatlimab and nivolumab lead-in arms, higher than previously reported, and conversely, lower (14%) in the nivolumab-relatlimab lead-in arm.

### Early pathologic response on biopsy is differentially impacted by lead-in therapy and serves as a surrogate for long-term disease outcome

Pathologic response assessed on biopsy including residual viable tumor (RVT), proliferative fibrosis, neovascularization, lymphoid aggregates, degree of tumor-infiltrating lymphocytes, and plasma cells were recorded for 35 evaluable patients who underwent serial biopsies of prespecified target lesions at baseline and week 4; pre and post tissue biopsy sites were identical in all patients (**Fig. S3A-G**). RVT was ≤10% in 48% (n=17) of cases. Proliferative fibrosis, neovascularization, plasma cells, moderate to high TIL, and lymphoid aggregates were identified in 49% (n=17), 40% (n=14), 40% (n=14), 54% (n=19), and 17% (n=6) of cases, respectively. Relatlimab lead-in therapy yielded a RVT≤10% in only 27% (n=3) cases, whereas nivolumab alone and the rela-nivo combination yielded 60% (n=6) and 57% (n=8) of cases with RVT≤10%, respectively **(Fig. S4A-F)**. All components of immune-related pathologic response at week 4 were less evident in biopsy samples from patients on the relatlimab lead-in arm **(Fig. 2A**). Major pathologic response on biopsy (MPRbx), which corresponds to immune-related pathologic response (irPR) score of 3 at week 4 assessment was 31.4% (n=11) in the whole cohort, and significantly lower with relatlimab-*vs.* nivolumab-(0% vs 50%, p=0.01) and relatlimab-*vs*. combination-(0% vs 43%, p=0.02) lead-in arms, respectively **(Fig. 2B**). No significant differences were observed between MPRbx and irPR of nivolumab *vs.* combination-treated samples. Amongst all samples, MPRbx was associated with early radiological response at week 4 (p=0.008). Patients with radiological findings of SD had irPR scores of 0=3, 1=7, 2=1, and 3=8, highlighting the heterogeneity of pathological response among patients with radiologically-defined stable disease at 4-week assessment (**Fig. 2C)**. MPRbx was significantly associated with overall best response (p=0.007), and improved PFS (HR=0.30, 95% CI, 0.11 to 0.82, p=0.02) **(Fig. 2D)**.

**Fig. 2:**
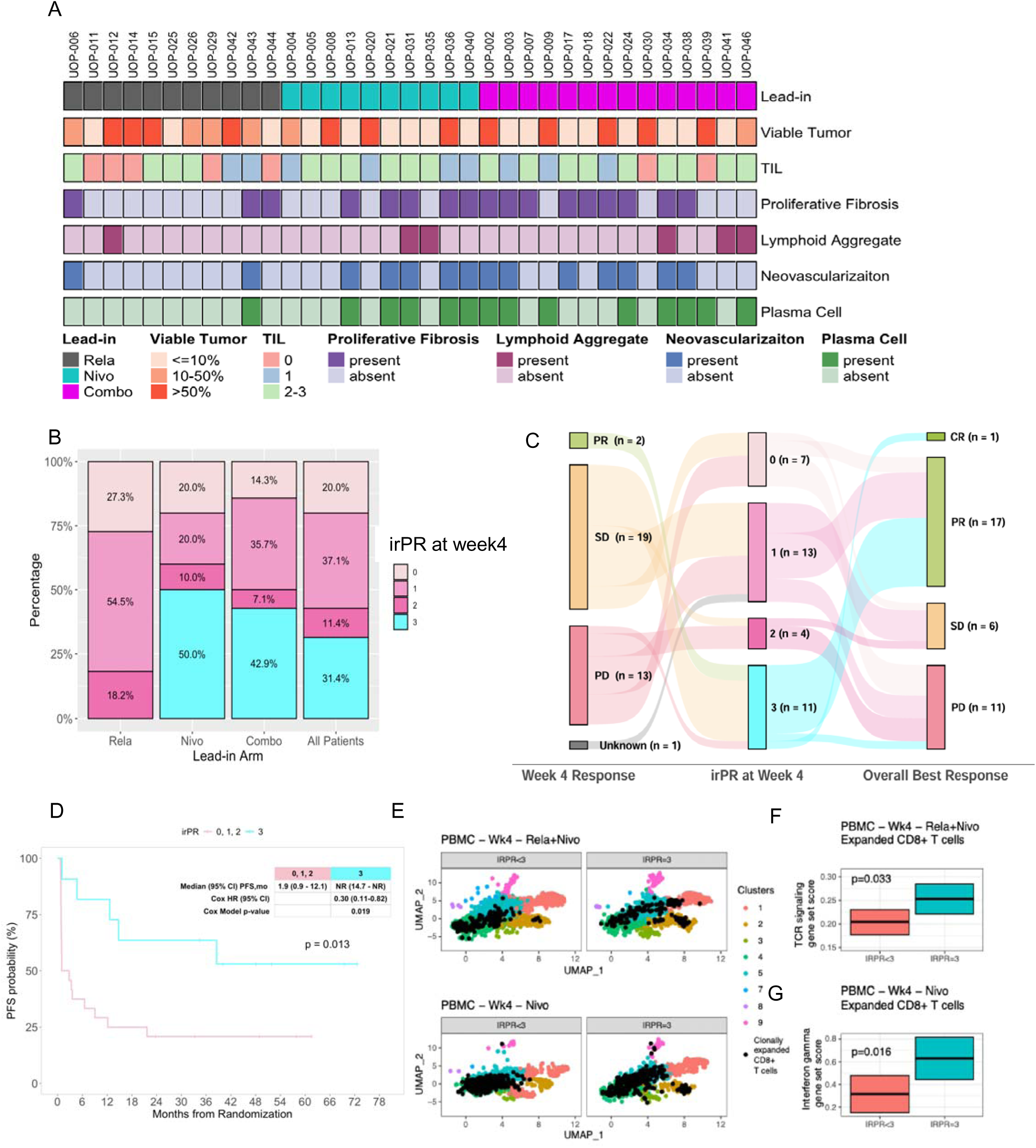
Major pathologic response on biopsy in relation to efficacy outcomes and peripheral blood immune changes. **(A)** Heat map of pathologic response features according to lead-in arm. (**B)** Bar plot showing proportion of immune-related pathologic response (irPR) 0, 1, 2, and 3 among all patients and per lead-in arm. (**C)** Sankey diagram showing irPR in relation to week 4 radiological response and overall best response. (**D)** Superior PFS (p=0.013, log-rank test) is associated with pathologic response (irPR=3) versus irPR equal to 0, 1 or 2 from week 4 biopsies, cox HR=0.30, [95% CI, 0.11 to 0.82, p=0.02]). (**E)** UMAPs of CD8+ T cells from peripheral blood of patients at week 4 post treatment. UMAPs of CD8+ T cells are shown from patients with IRPR<3 for Rela+Nivo and Nivo only treatment and for IRPR=3. Cluster assignments are derived from Cillo et al, Cell 2024. Clonally expanded T cells (that is, CD8+ T cells with TCRbeta CDR3 counts greater than 3) are denoted as black dots in their positions on the UMAP. (**F)** Mixed linear effects model revealed significantly increased TCR signaling transcriptional signatures in clonally expanded CD8+ T cells from IRPR<3 and IRPR=3 4 weeks following relatlimab+nivolumab (Rela+Nivo). (**G)** Mixed linear effects model revealed significantly increased interferon gamma response signatures in clonally expanded CD8+ T cells from IRPR<3 and IRPR=3 4 weeks following nivolumab (Nivo).

In order to assess systemic immune changes in relation to MPRbx for nivolumab and combination lead-in arms, we evaluated our previously identified gene signature scores in clonally expanded CD8^+^ T cells in PBMC at week 4 after each immunotherapy regimen **(Fig. 2E)**.^10^ Gene sets associated with TCR signaling in CD8+ T cells were associated with MPRbx (irPR=3) following combination therapy and gene sets associated with interferon gamma signaling in CD8+ T cells were associated with MPRbx after nivolumab monotherapy (**Fig. 2F, G**). These findings demonstrate that relatlimab *vs.* nivolumab or combination therapy resulted in lower pathologic response rates at 4-week assessment, and that MPRbx at 4 weeks among patients with advanced melanoma on first-line treatment is a useful surrogate for long-term improved disease outcome and associated with distinct peripheral blood immune changes that are differentially driven by lead-in nivolumab *vs.* combination therapy.

### Differential modulation of tumor immune microenvironment by lead-in therapy impacts treatment outcome

We further evaluated changes in the immune microenvironment induced by the three lead-in arms, and their relation to clinical outcomes. This was accomplished not only through assessment of immune cell densities, but also their spatial distribution in relation to tumor, and among one another within viable tumor, tumor bed, and peritumoral area. Considering high expression of *LAG3* on CD8^+^ T cells, we first assessed the impact of lead-in therapy on tumor immune microenvironment changes of CD8^+^ T cells in relation to other immune subtypes, using a custom multiplex immunofluorescence panel (**Fig. 3A, B**). Combination nivolumab-relatlimab and single agent nivolumab, but not relatlimab lead-in treatment resulted in significant increases in the density of CD8^+^ T cells (p<0.01 and p=0.01, respectively) (**Fig. S5A, B**). MPRbx correlated with a significantly higher intratumoral CD8⁺ T-cell density (**Fig. S5C**). The density of CD8^+^ T cells varied amongst on-treatment melanoma samples and was divided into quartiles. On-treatment week 4 but not baseline CD8^+^ T cell density trended toward association with improved PFS, with highest PFS observed in tumors with highest quartile CD8^+^ T cell density (**Fig. S5D**). There was no significant correlation between CD8^+^ T cells and conventional T cells or T_regs_ at baseline or week 4. Baseline but not week 4 CD8^+^ T cells were correlated with CD20^+^ B cells (r=0.37, p=0.037) and CD68^+^ macrophages (r=0.41, p=0.02) (**Fig. S5E, F**). We next evaluated CD8^+^ T cell clustering patterns with tumor and other immune subtypes. While were was no significant correlation between SOX10^+^ tumor cells and CD8^+^ T cells in baseline biopsy samples, spatial co-clustering analysis revealed that baseline tumors in the relatlimab lead-in arm had significant increased dispersion of SOX10^+^ tumor cells and CD8^+^ T cells (p=0.009 and p=0.07 in comparison to combination and nivolumab lead-in arms, respectively) indicating separation of the patterns and low colocalization of signals (**Fig. 3C**). Notably, CD8^+^ T cell and SOX10^+^ tumor cell colocalization was associated with improved PFS indicating the importance of tumor-immune cell interaction for effective anti-tumor activity **(Fig. 3D)**. Further assessment of baseline spatial co-clustering of immune cells revealed that relatlimab alone was characterized by significant clustering of CD8^+^ with T_regs_ within tumor regions (p=0.026 and p=0.01 compared nivolumab and combination lead-in arms, respectively) with CD8^+^ and FOXP3^+^ T_reg_ clustering associated with worse PFS (p=0.018) (**Fig. 3 E, F**). CD8⁺–FOXP3⁺ cell clustering significantly influenced progression-free survival, independent of CD8⁺ T-cell density, underscoring the relevance of spatial immune colocalization within the tumor microenvironment (**Fig. 3G**). To evaluate whether this clustering of CD8^+^ and T_regs_ continued after the lead-in interval we reviewed week 4 samples, which strikingly revealed that the relatlimab lead-in arm continued to be characterized by significantly increased clustering of CD8^+^ and T_regs_ (p=0.022 and p=0.047 in comparison to nivolumab alone and combination lead-in arms, respectively (**Fig. 3H**). In order to further interrogate interplay between CD8^+^ and FOXP3^+^ T_regs_, we performed cell-cell interaction analysis using single cell RNA sequencing data obtained at baseline and week 4 across the three lead-in arms **(Fig. S6A)**. We first generated circos plots to visualize interactions at baseline, before initiation of any therapy, and at week 4 for relatlimab, nivolumab, and combination lead-in arms **(Fig. S6B-E)**. We next calculated interaction ratios to identify top enriched ligand-receptor interactions in relatlimab lead-in arm in comparison to other treatment arms **(Fig. 3I)**. Immune checkpoint-mediated interactions upregulated in post-relatlimab therapy included *CD80/CTLA4, ICOSLG/CTLA4, SIGLEC7/TIMD4, TIGIT/SIRPA, NECTIN3/TIGIT, NECTIN2/TIGIT, TIGIT/RECK, CD96/NECTIN2* and *CD274/PDCD1*.

**Fig. 3:**
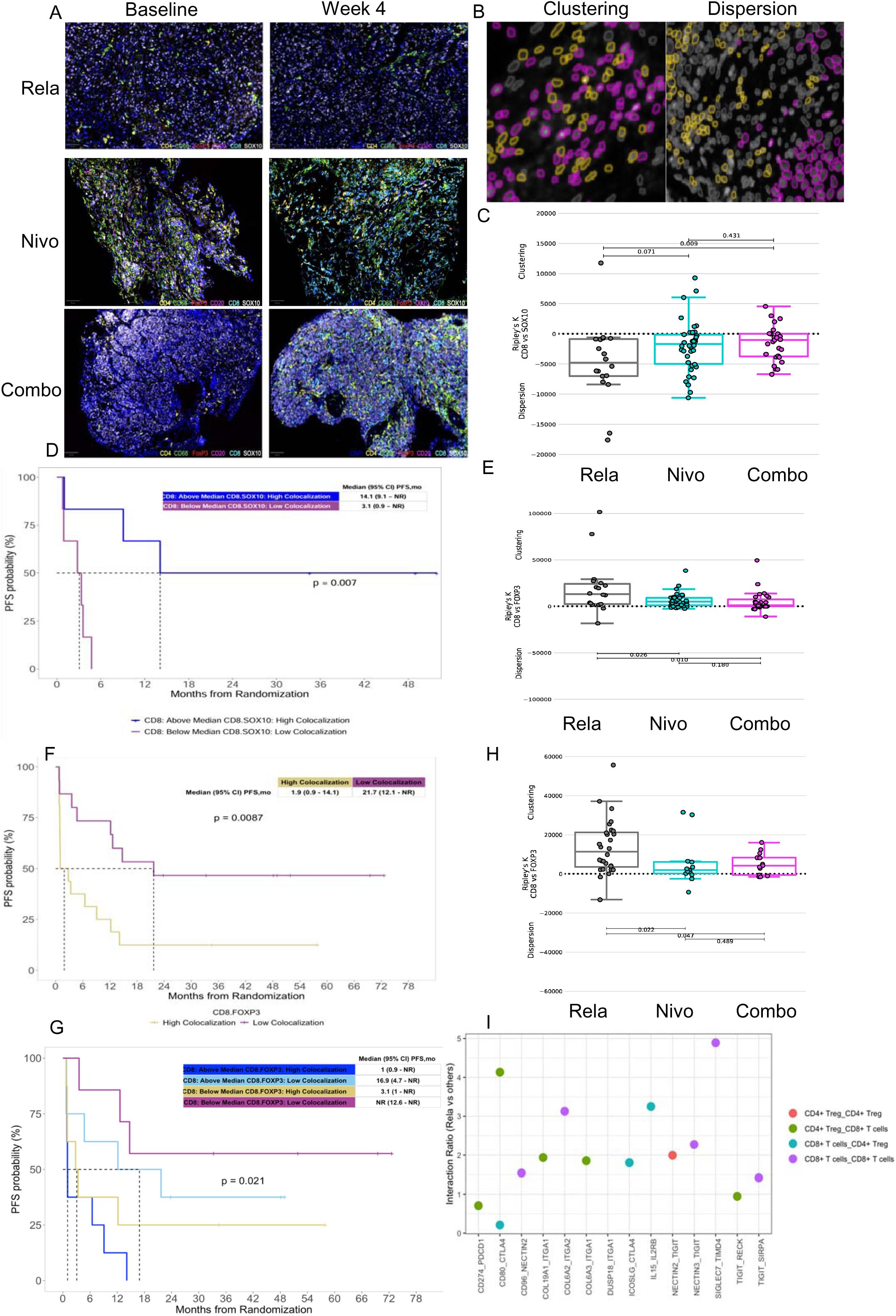
Lead-in therapy induced immune changes in tumor microenvironment (TME) in relation to anti-tumor efficacy outcomes. **(A)** Representative multispectral Immunofluorescence image of a melanoma TME. Tissues were stained with the following primary antibody/conjugated opal pairs: CD8/Opal480, CD4/Opal690, CD68/Opal520, SOX10/Opal780, FOXP3/Opal570, CD20/Opal620. Each tissue image was segmented into individual cells using the DAPI marker for cell nuclei. (**B)** Image illustrates clustering (left) and dispersion (right) of two distinct immune subtypes (yellow and purple). (**C)** Box plot indicates baseline colocalization patterns of CD8^+^ T cells and SOX10^+^ tumor regions. Ripley’s K function is illustrated on y axis. Higher numbers indicate clustering and lower numbers indicate dispersion of these two cell types. The comparison of baseline samples among 3 lead-in arms reveals significant dispersion of CD8^+^ T cells from tumor cells in relatlimab lead-in arm vs combination indicating low tumor-CD8^+^ T cell colocalization. P values were calculated using a Wilcoxon rank-sum test. (**D)** PFS according to CD8^+^ T cell density and CD8^+^ T cell and SOX10 tumor cell colocalization. (**E)** Box plot indicates baseline colocalization patterns of CD8^+^ T cells and FOXP3^+^ T regulatory cells. Ripley’s K function is illustrated on y axis. Higher numbers indicate clustering and lower numbers indicate dispersion of these two cell types. The comparison of baseline samples among 3 lead-in arms reveals significant clustering of CD8^+^ T cells and FOXP3^+^ T regulatory cells in relatlimab lead-in arm vs nivolumab and combination indicating high FOXP3^+^ T regulatory -CD8^+^ T cell colocalization. P values were calculated using a Wilcoxon rank-sum test. (**F)** PFS according to FOXP3^+^ T regulatory -CD8^+^ T cells colocalization. (**G)** CD8^+^ T cells and FOXP3^+^ T regulatory cell colocalization at week 4. (**H)** Top enriched ligand-receptor pairs in rela lead-in arm in comparison to other treatment arms. Dot plot to show communicating cell subtypes (by color), interaction ratio (y axis) and ligand-receptor pairs (x axis).

Regarding the impact of lead-in therapy on other immune cell subtypes, no significant differences were observed among co-clustering patterns of CD8^+^ T cells and CD20^+^ B cells, or CD68^+^ macrophages, among the three lead-in arms at baseline and subsequently on-treatment (**Fig. S7 A-D**). Combination lead-in therapy, but not nivolumab or relatlimab lead-in therapies also significantly increased CD4^+^FOXP3^-^ T cells (p=0.03) but did not significantly impact the density of CD20^+^ B cells or CD68^+^ macrophages (**Fig. S7 E-G**).

Our findings demonstrate that the melanoma TME at baseline is characterized by a wide range of CD8^+^ T cell infiltration densities. While simple correlative analysis of CD8^+^ T cells and SOX10^+^ tumor cells did not reveal significant changes, it is notable that those two populations were mainly dispersed in the TME at baseline. Baseline tumor CD8^+^ count did not demonstrate any association with PFS, but co-clustering of CD8^+^ T cells with FOXP3^+^ T_regs_ was associated with poor disease outcome. This finding suggests that the distribution of immune cells in the TME may predict treatment outcome, even in cases where cell density does not predict immunotherapy response. Evaluation of TME changes induced by lead-in therapy revealed that nivolumab and combination therapy increased total CD8^+^ T cell counts, and post-treatment high CD8^+^ T cell populations were associated with improved PFS. In contrast, relatlimab lead-in exposure resulted in co-clustering of CD8^+^ and FOXP3^+^ T_regs_, up-regulation of CTLA4-and TIGIT-mediated interactions, potentially adversely impacting the clinical outcome in this cohort.

### Peripheral blood CD8^+^ T cell responses following lead-in therapy

Considering CD8^+^ T cells and their subsets in peripheral blood are the main subpopulations of PBMC that express *LAG3*, we asked whether those were differentially modulated by lead-in therapy in this trial. Lead-in therapy with nivolumab, relatlimab or the combination did not modulate total CD8^+^ T cell counts between baseline and week 4 assessment (**Fig. 4A**). However, the combination, but neither nivolumab nor relatlimab lead-in therapy alone, resulted in significant increases in the CD8^+^ effector memory (T_EM_) subset, without significant changes in CD8^+^ central memory (T_CM_) or naive CD8^+^ T cell subsets (**Fig. 4B-D**). We next sought to evaluate whether any of these changes were associated with clinical outcomes. Patients with objective clinical response demonstrated a significant increase in the CD8^+^ T_EM_ subset (**Fig. 4E**). Next, we assessed Ki67^+^CD8^+^ populations considering their previously described correlations with immune response ^11,12^. Relatlimab, in comparison to other lead-in arms showed a trend to increased Ki67^+^CD8^+^ subsets. Moreover, patients with progressive disease showed an increase in Ki67^+^CD8^+^ subsets **(Fig. 4F-G)**. Further classification of those Ki67^+^CD8^+^ T cells revealed an increase in Ki67^+^CD8^+^ T_EM_ and Ki67^+^CD8^+^ T_CM_ cells in patients with progressive disease while the relatlimab arm specifically showed increased Ki67^+^CD8^+^ T_CM_ cells **(Fig. 4H-K)**. The data underscores the importance of CD8^+^ T_EM_ subsets which were significantly increased in combination lead-in arm and associated with overall response to therapy.

**Fig. 4:**
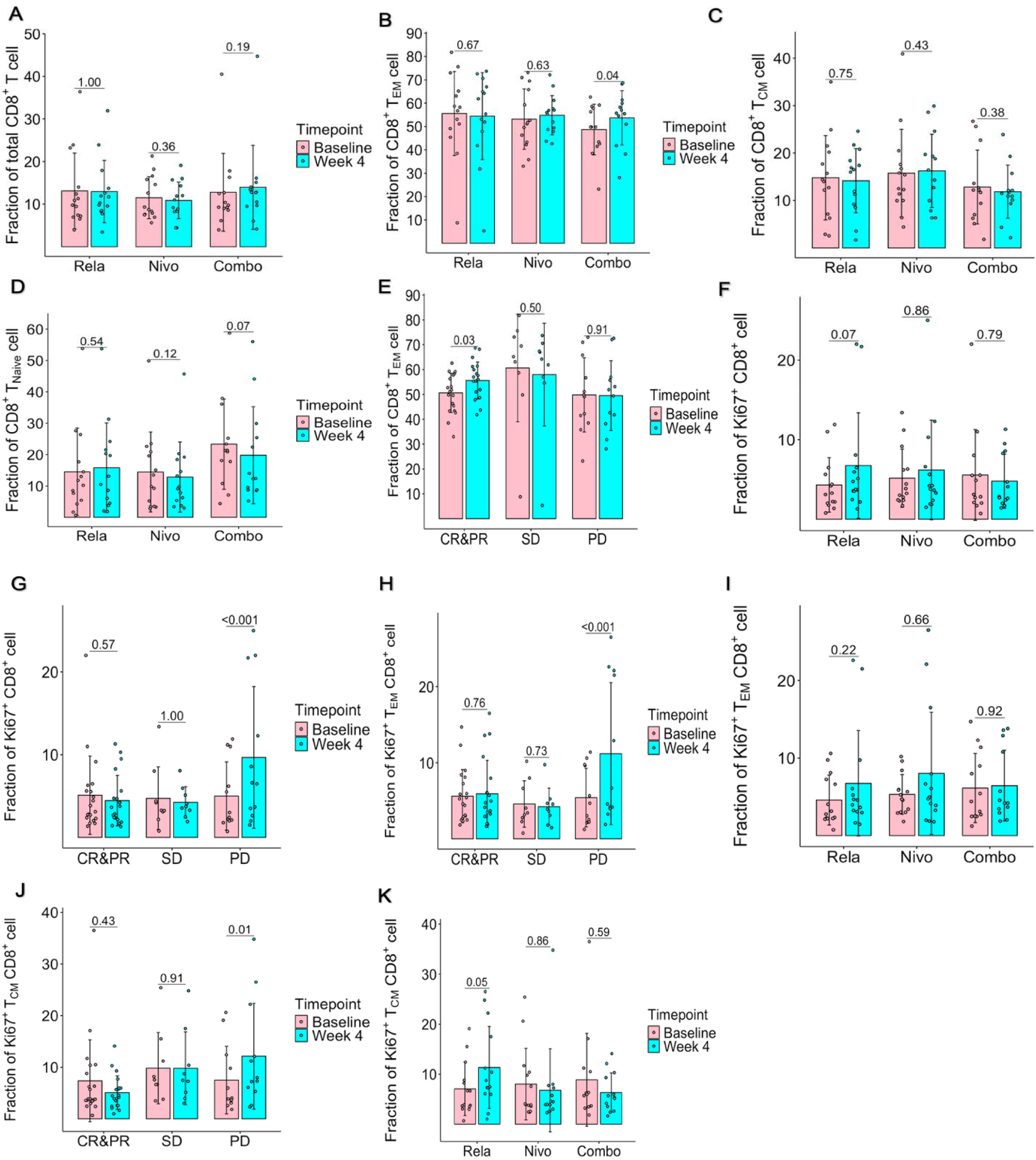
Lead-in therapy induced changes in CD8^+^ T cells in association with efficacy outcomes in peripheral blood. **(A)** Lead-in therapy induced changes in total CD8^+^ T cells. No significant differences were observed in total CD8^+^ T cell population among 3 lead-in arms. (**B)** Lead-in therapy induced changes in CD8^+^T effector memory cells. Combination (Combo) therapy significantly increased CD8^+^T effector memory cells**. (C)** Lead-in therapy induced no significant changes in central memory (T_CM_) CD8^+^ subsets. (**D)** Lead-in therapy induced no significant changes in CD8^+^T_NAIVE_ subsets. Wilcoxon signed-rank test p value is depicted on the figure per lead-in arm. (**E)** Response to therapy is associated with increase in CD8^+^T effector memory cells. (**F)** Lead-in relatlimab trended to increase K67^+^CD8^+^ T cells. (**G)** Progressive disease was associated with increase in total Ki67^+^CD8^+^ T cells. (**H)** Progressive disease was associated with increase in total Ki67^+^CD8^+^ effector memory T cells. (**I)** Lead-in therapy induced changes in Ki67^+^CD8^+^T effector memory cells. (**J)** Progressive disease was associated with increase in total Ki67^+^CD8^+^ T_CM_ cells. (**K)** Lead-in relatlimab increased K67^+^CD8^+^ T_CM_ cells. Abbreviations are Rela: relatlimab, Nivo: nivolumab, Combo: combination. CR&PR: complete and partial response, SD-stable disease, PD-progressive disease

### Decrease in CD33^dim^ monocytic populations in peripheral blood impacts overall disease outcome

Given the clinically meaningful PFS differences among patients who received relatlimab lead-in followed by combination therapy, we used an unbiased approach to assess all immune subsets in peripheral blood using high dimensional multispectral flow cytometry to decipher whether unique immune subsets are differentially impacted by relatlimab, nivolumab, or the combination in relation to overall disease outcome. We first used t-SNE to gate on live CD45^+^ cells. FlowSOM clustering algorithms identified 17 metaclusters, among which metacluster 14 was decreased in relatlimab lead-in arm patients at week 4.

Manual gating overlay identified those as subpopulations of CD14^++^CD16^−^ classical monocytes (**Fig. 5A**). Interestingly, this same population was decreased in patients with progressive disease. **(Fig. 5B)**. Further subclustering of CD14^+^ myeloid cells highlighted their median intensity of HLA-DR, CD33, CD38 and other phenotypic markers **(Fig. 5C)**.

**Fig. 5:**
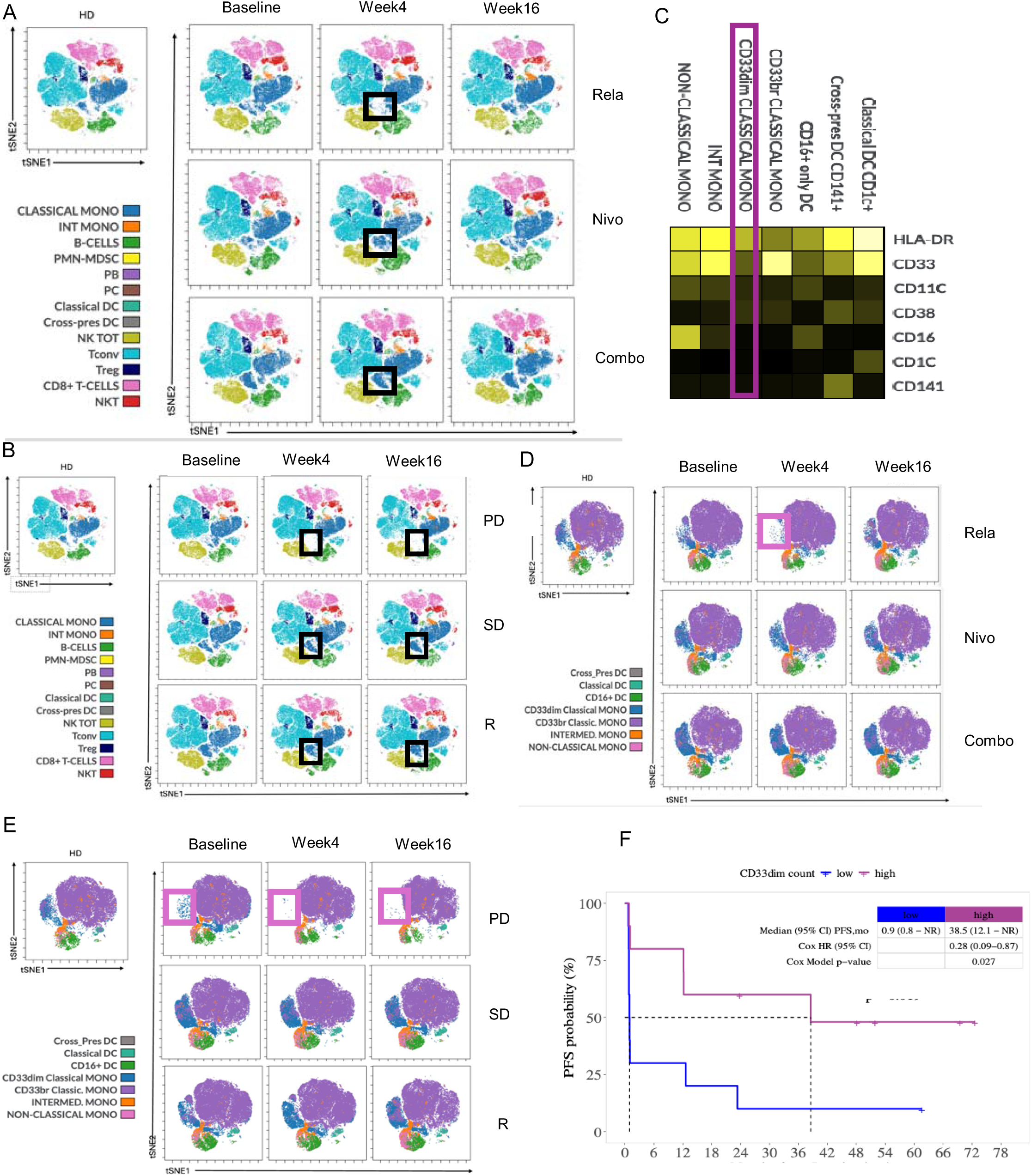
Lead-in therapy induced peripheral blood immunophenotyping in association with anti-tumor efficacy outcomes. **(A)** t-SNE algorithm from total CD45^+^ population, immune cell subpopulations were manually gated. Relatlimab (Rela) lead-in arm was characterized by decrease in subpopulations of classical monocytes. (**B)** Patients with progressive disease (PD) were characterized with baseline diminished subclusters among classical monocytes which were further decreased at week 4 and week 16. Black square highlights diminished subpopulations of classical monocytes. (**C)** Heat map shows mean expression of 7 markers. Heat colors show overall expression levels (yellow, high expression; dark black, no expression). (**D)** t-SNE algorithm from total CD14^+^ population, myeloid cells were manually gated. Relatlimab (Rela) lead-in arm was characterized by decrease in CD33^dim^ classical monocytes. (**E)** Patients with progressive disease (PD) were characterized by baseline diminished CD33^dim^ classical monocytes, which were further decreased at week 4 and week 16. Pink square highlights diminished subpopulations of classical monocytes. (**F)** Progression-free survival according to CD33^dim^ classical monocytes.

We further characterized CD33^dim^ monocytes subset using scRNA sequencing data derived from PBMCs. Leiden-based clustering identified 9 clusters in myeloid cells (clusters 0 through 9). Clusters 0, 1, and 9 were classical monocytes (**Fig. S8A**). We assessed differentially expressed genes across these 3 clusters and revealed that cluster 1 (CD33^dim^) was transcriptionally distinct from cluster 9 and shared some features with cluster 0 (**Fig. S8B**). Using curated gene sets from the Molecular Signatures Database cluster 1 showed enrichment in IFN alpha response (similar to cluster 0) and NOTCH signaling with downregulation of gene sets in IL6_JAK_STAT3 signaling, epithelial mesenchymal transition, and TGF beta signaling pathways (**Fig. S8C**). We next assessed clinical impact of this CD14^+^CD16^-^HLA-DR^low^CD33^dim^ metacluster showing that it was both under-represented at baseline and diminished at weeks 4 and 16 among patients with progressive disease in association with worse PFS **(Fig. 5D, E, F)**. These data indicate that the CD14^+^CD16^-^ HLA-DR^low^CD33^dim^ subcluster was diminished in patients with progressive disease, warranting further investigation of this subpopulation of monocytes in relation to anti-tumor activity.

## Discussion

Treatment targeting co-inhibitory immune checkpoint molecules including PD-1, CTLA-4, and LAG-3 have become the cornerstones of management for melanoma, and other solid tumors.^13^ Combined checkpoint blockade has been the most effective therapeutic strategy for advanced melanoma for nearly a decade, and has recently entered the neoadjuvant setting. Historically anti-PD-1, as the most active single agent, has been administered with anti-CTLA-4 or anti-LAG-3, and anti-PD1 has been the standard of care in the adjuvant setting.^14–16^

The current study investigated the clinical efficacy of the combination nivolumab-relatlimab in relation to its component single agents. To do this, we developed a novel four-week lead-in treatment in which consenting patients were assigned at random to receive nivolumab, relatlimab, or nivolumab-relatlimab therapy. We leveraged this unique trial design and biospecimens (tumor tissue and peripheral blood samples) to assess immunological changes associated with the component therapies. Although this study was conceived as an immune correlate study and was not powered to detect differences in clinical outcomes between lead-in treatment arms, we nevertheless found that in intention-to-treat analysis of median PFS was 6.5 months, which was lower than reported in the pivotal phase 3 trial. A major driver of this difference was the short median PFS of only 1.9 months in the relatlimab lead-in arm (compared with 4.7 months in the nivolumab lead-in arm and 14.7 months in the combination lead-in arm). Similarly, ORR was 21% in the relatlimab lead-in arm, and ∼60% for nivolumab and for the combination lead-in arms. Despite comparable early response data, including MPRbx and best overall response, the long-term outcomes between the nivolumab and combination lead-in arms diverged substantially. The 4-year PFS rate was markedly lower with nivolumab monotherapy, and its median PFS closely mirrored historical data for single-agent anti–PD-1 therapy.^2,6^ Acquired resistance to anti–PD-1 therapy has been increasingly recognized, with several mechanistic pathways implicated. These include defects in antigen presentation driven by loss-of-function mutations in β2-microglobulin, impaired interferon receptor signaling pathways, and compensatory upregulation of alternative immune checkpoints such as LAG-3 and TIM-3.^17–19^ Thus, this short lead-in therapy with relatlimab or nivolumab alone unexpectedly seemed to influence outcome.

Pathologic assessment of response after neoadjuvant therapy represents a well-established endpoint for assessment of immunotherapy efficacy. In a pooled efficacy analysis of neoadjuvant trials for Stage III/IV melanoma, pathologic response was correlated with long-term RFS and OS. Pathologic response assessed early (after 2-3 cycles of immunotherapy) served as a reliable surrogate marker for long-term efficacy.^20,21^ Early assessment of antitumor activity has been performed through simple histological assessment of pathologic response in the adjuvant/neoadjuvant setting, but has not previously been probed in the setting of first-line therapy of metastatic melanoma. For the assessment of patients with advanced inoperable disease, full resection of tumor is not feasible, and we have utilized core biopsies for tumor assessment. Immune-related pathologic response assessment upon biopsy material has previously been studied in metastatic melanoma patients receiving anti-PD-1 therapy.^22^ This study has shown that MPRbx is associated with later radiological response and ultimately with overall survival. Core biopsies represent part of the tissue, in contrast to surgical excisions that comprise the whole tumor, it is important to note that immune-related pathologic response assessment measures residual viable tumor percentage, and takes into consideration tissue proliferation, and immune activation markers. In cases where RVT is <10% but lacks other stipulated markers of response, the assessment does not classify a particular patient as a pathological responder. In this study we assessed the impact of lead-in therapy through prospective evaluation of MPRbx at week 4. Pathologic response assessment was available in a majority (81%) of cases with data lacking in the remainder due to early clinical progression, death due to adverse events, or lesion disappearance (antitumor response). MPRbx was recorded at week 4 of therapy was overall 31.4% and varied among the different lead-in arms with 0% on relatlimab lead-in, while it was 50% and 42% for the nivolumab-and combination-lead-in recipients, respectively. MPRbx at week 4 was associated with later best overall radiological response and with improved PFS, showing that MPRbx is an early biomarker for immunotherapy efficacy in advanced melanoma. Notably, MPRbx was associated with our previously reported nivolumab-induced interferon gamma signaling-and combination-induced TCR signaling-gene sets in CD8^+^ T cells emphasizing biological impact of early pathologic response and differences in nivolumab versus relatlimab+nivolumab lead in arms.^10^

Enhanced co-clustering of CD8^+^ T cells and FOXP3^+^ T_regs_ in relatlimab lead in arm was characterized with pathways enriched in immune checkpoints which would require further mechanistic investigation. There are ongoing clinical trials in the first-line advanced melanoma setting of combining nivolumab-relatlimab with ipilimumab which demonstrated encouraging efficacy data in preliminary reports along with improvement of safety with addition of IL-6R inhibitor.^23,24^

Previous studies have identified peripheral blood T cell and myeloid compartment changes in relation to response to anti-PD-1 therapy in melanoma. In our study, peripheral blood immunophenotyping evaluation of serial samples revealed distinct changes driven by the separate lead-in arms.^25^ Combination therapy led to an increase in CD8^+^ T_EM_ subsets that were associated with overall response to therapy. Elevated baseline CD8^+^ T_EM_ cells have been associated with improved OS and clinical response to ipilimumab therapy in patients with melanoma.^26^ Notably, in our study Ki67^+^CD8^+^ T cell subsets were increased in progressive disease patients consistent with previous results which showed that Ki67^+^CD8^+^ T cell subsets increased in melanoma versus healthy control PBMCs, positively correlated with tumor burden in relation to poor prognosis.^12^ Monocytes have been described in general as immunosuppressive populations and in one previous neoadjuvant nivolumab-relatlimab study tumor CD68^+^HLA-DR^+^CD14^+^VISTA^+^CD163^+^CD45RO^+^ PD-L1^+^ macrophages were associated with non-response to therapy.^14^ The CD14^+^CD16^-^HLA-DR^low^CD33^dim^ subcluster was decreased among patients with progressive disease, both at baseline and on-treatment. CD14^+^CD16^−^CD33^+^HLA-DR^hi^ monocytes were found to predict response to anti-PD-1 therapy in melanoma, supporting the findings that this subpopulation of monocytes has a role in anti-tumor activity and to those potential mechanisms of therapeutic efficacy, and their analysis as mechanisms of resistance.^27^

In summary, first-line nivolumab-relatlimab combination therapy of advanced melanoma has been dissected in this novel study using lead-in monotherapy with relatlimab, nivolumab or the combination. We have noted diminished efficacy of nivolumab and relatlimab given as a lead-in monotherapy, despite receipt of the combination subsequent to week 4. These findings warrant consideration in clinical practice. The novel analysis of component agents afforded by this novel trial design has identified different immunological mechanisms of anti-PD1 and anti-LAG3 in comparison to the combination, and in relation to MPRbx which serves as an early surrogate marker for immunotherapy response and long-term PFS. The combination resulted in durable responses with acceptable safety confirming the efficacy of the nivolumab+relatlimab combination given as first line therapy for advanced melanoma patients.

## Materials and Methods

### Patients

Patients were eligible if they were at least 18 years old, ECOG performance status ≤1 with AJCC 8th edition criteria for unresectable stage III or stage IV melanoma who have not received prior immune checkpoint blockade treatment in the metastatic setting. Patients who had received prior adjuvant treatment with anti-PD1, anti-PDL1, and/or anti-LAG3 antibody were excluded. Prior adjuvant treatment with targeted therapy, anti-CTLA4, or treatment not otherwise specified was permitted. Patients were required to have normal organ and marrow function including leukocytes ≥3,000/mcL, absolute neutrophil count ≥1,500/mcL, platelets ≥100,000/mcL, total bilirubin ≤1.5 × institutional upper limit of normal (except subjects with Gilbert’s syndrome), AST(SGOT)/ALT(SGPT) ≤2.5 × institutional upper limit of normal, creatinine within normal institutional limits or creatinine clearance ≥60 mL/min/1.73 m2 for patients with creatinine levels above institutional normal. Subjects must have presence of measurable disease pre RECIST v1.1. Patients with known or suspected active CNS metastases were excluded unless they underwent surgical or radiation treatment and demonstrated no progression of disease at week 4 radiological assessment after the initial definitive therapy and required no steroid therapy. Subjects receiving any other investigational agents, allergic reactions attributed to compounds similar to nivolumab or relatlimab, history of life-threatening toxicity related to prior immune therapy, uncontrolled or significant cardiovascular disease, confirmed history of encephalitis, meningitis, or uncontrolled seizures in the year prior to informed consent were excluded. Subjects who had a known additional malignancy that is progressing or requires active treatment were excluded unless basal cell carcinoma of the skin or squamous cell carcinoma of the skin that has undergone potentially curative therapy or *in situ* cervical cancer. Sex or gender was self-reported, and patients were enrolled regardless of sex or gender. Pregnant women were excluded due to unknown safety of relatlimab in this population. Subjects with active, known or suspected autoimmune diseases were excluded with exceptions of type I diabetes mellitus, hypothyroidism only requiring hormone replacement, skin disorders (such as vitiligo, psoriasis, or alopecia) not requiring systemic treatment. Baseline characteristics of patients are presented in Table 1 and full details of exclusion/inclusion criteria are reported in the study protocol in the supplementary information.

### Study Design and Treatment

This was a single center phase 2 study designed to evaluate the change in immune biomarkers of treatment with 4 weeks of lead-in relatlimab alone, nivolumab alone, or the combination of nivolumab and relatlimab in relation to objective response rate to the combination of nivolumab/relatlimab as compared to historical studies of patients with unresectable or metastatic melanoma who have not received prior treatment with immunotherapy for metastatic disease. In the lead-in phase, subjects were randomized 1:1:1 to one of three lead-in treatments: nivolumab alone (480 mg IV once), relatlimab alone (160 mg IV once), or the combination therapy (Nivolumab administered at 480 mg IV followed by infusion of relatlimab at 160 mg IV) for 1 cycle (4 weeks). Following completion of 1 cycle of assigned lead-in treatment, all subjects were reassessed clinically and by staging CT imaging, and then would proceed to the combination for treatment every 4 weeks with nivolumab + relatlimab until progression or dose-limiting toxicity. Maximum planned duration of treatment was 24 months (24 cycles). CT imaging performed for disease assessment was performed at 12 weeks (+/-1 week) following initiation of combination phase, and then at 12-week (+/-1 week) intervals throughout treatment.

### Statistical Methods

The primary objectives of the clinical trial were to (1) assess changes in immune biomarkers and estimate the association between changes in immune biomarkers and changes in tumor size during the lead-in phase, and (2) to access the objective response rate relative to the beginning of the combination phase by comparing CT measurements obtained 12 weeks (+/-1 week) after beginning of combination phase and subsequent 12 week (+/-1 week) intervals to CT measurements obtained at initial 4 week completion of lead-in phase. Secondary objectives included evaluating (1) the clinical benefit of the combination therapy, (2) duration of response (DOR), (3) progression-free survival (PFS), (4) overall survival (OS), (5) the safety of Nivolumab and Relatlimab, and (6) changes in immune biomarkers. Exploratory objectives included the assessment of pathological response at week 4.

The sample size calculation for this trial was based on an expected clinical response rate of 40% with Nivolumab in treatment-naïve patients with unresectable stage IIIB, stage IIIC, or stage IV melanoma, and an anticipated 60% with the addition of Relatlimab. A single-arm, one-stage design with α = 0.10 has been selected, requiring 41 patients to provide 90% power for a one-sample, one-tailed exact binomial test. Since patients were to be randomly assigned to one of three lead-in regimens, 42 response-evaluable patients would be accrued and complete 12 weeks of treatment, allowing for exactly 14 patients per lead-in group. Due to the development of antitumor response at week 4 in one patient who had lethal cardiac adverse immune toxicity one additional patient was accrued for a total enrollment of 43 patients.

In this manuscript our report focuses on the primary endpoint of the objective response rate (ORR, defined as the sum of complete and partial responses divided by the total number of patients assessed) along with exact 90% confidence intervals using the Clopper-Pearson Method, which was initially assessed at 12 weeks after the initiation of the combination phase (week 16) compared to baseline imaging obtained immediately prior to initiating combination (week 4). Analyses were performed in both the intention-to-treat population, which included all randomized patients (n=43 ; patients without available response data were considered non-responders), and the evaluable population.

For the secondary objectives, clinical benefit rate was defined as the sum of complete responses, partial responses, and stable disease divided by the total number of patients, calculated along with 90% confidence intervals, initially assessed at 12 weeks after the initiation of the combination phase and compared to baseline imaging obtained immediately prior to starting the combination therapy. All subjects who received at least one dose of Relatlimab, Nivolumab, or the combination were analyzed for safety, and adverse events were tabulated.

Safety was measured in terms of adverse events that were graded using the Cancer Therapy Evaluation Program (CTEP) Common Terminology Criteria for Adverse Events (CTCAE) version 5.0. Duration of response (DOR) was calculated from the time measurement criteria were first met for complete response or partial response (whichever is first recorded) until the first date that progressive disease was objectively documented or death. DOR was assessed only among patients who achieved a complete or partial response. Progression-free survival (PFS) was calculated as the time from the date of randomization to the first date of documented progression or death due to any cause, whichever occurred first. Overall survival (OS) was calculated as the time from the date of randomization to the date of death due to any cause.

Survival endpoints (OS, PFS, and DOR) were summarized and plotted with the Kaplan-Meier method, and compared between subgroups, defined by lead-in arms, BRAF mutation status, gender, and week 4 pathological response etc., using the log-rank test. Their association with explanatory variables, such as immune cell density etc., were studied with univariable and multivariable Cox proportional hazards models.

Best overall response (BOR) was defined as the best response recorded from treatment initiation. The best overall response rate (BORR), defined as the proportion of patients achieving a complete or partial response as their best overall response from treatment initiation, was reported with exact 90% Clopper–Pearson confidence intervals in the intention-to-treat population as well as in selected subpopulations of research interest.

For immune data, the densities of each immune cell type (4+FOXP3+, SOX10+, CD68+, CD4+FOXP3-, CD4, CD8, CD20, CD68, SOX10) were compared between baseline, week 4, and week 16 within each lead-in arm using the Wilcoxon signed-rank test. And their association with PFS was studied with univariable Cox proportional hazard models. For CD8 and CD20 cell density, a linear mixed-effects model which took into account patient heterogeneity was conducted to assess the impact of the lead-in arm over time. Associations between CD8 density, clustering value with CD8 and other cell types (CD4, CD20, CD68, FOXP3, SOX10), and PFS were investigated at baseline and week 4 within different tissue types. Multivariable associations of cell densities (CD4+FOXP3+, CD8, CD20, and CD68) at different time points with PFS were conducted. Non-parametric Spearman correlation test was used to assess the relationship between different immune cell densities at various time points. Peripheral blood immune biomarkers (such as CD45, CD14, and Treg) within each immune cell subgroup (including General population, MAC_MONO subpopulations, T memory populations, NK subpopulations, Cycling T cell populations, DC subpopulations, and small populations) were compared between baseline, week 4, and week 16 in each lead-in arm, as well as by best overall response, using the Wilcoxon signed-rank test. A linear mixed-effects model was conducted to detect the effects of time, lead-in arm, and best overall response as predictors for peripheral blood immune biomarkers from baseline to weeks 4 and 16.

For exploratory objectives, pathological response on biopsy was assessed after one cycle of lead-in treatment. Fisher’s exact test was used to evaluate associations between lead-in arms and pathological response, week 4 clinical response and pathological response, week 16 clinical response and pathological response, and best overall response and pathological response. Association between pathological response and PFS were also analyzed. All the above analyses were performed with R software (version 4.3.1).

### Pathologic response assessment

Pathologic response was assessed on a biopsy specimen performed based on the report of *Stein et al*. utilizing H&E-stained formalin-fixed and paraffin-embedded (FFPE) tissues that were reviewed by a single reference board-certified dermatopathologist (AK). The features such as percent residual viable tumor (RVT), proliferative fibrosis, neovascularization, plasma cells, lymphoid aggregates, and degree of tumor-infiltrating lymphocytes (absent, low, moderate or high) were recorded. Immune-related pathologic response (irPR) was classified 0 when no features were present, irPR=1 in cases with <3 features, irPR=2 with at least 3 colocalized features, where there was more than 10% RVT, and irPR=3 where there was evidence of major pathologic response (MPRbx) including at least 3 of the foregoing features along with ≤10% RVT.

### Multiplex Immunofluorescence Analysis

Individual antibodies were optimized as single stains according to supplier recommendations using appropriate control tissues. Multiplex panel optimization was carried out according to Akoya Bioscience’s “Opal Assay Development Guide” provided on the website. Automated multiplex staining of tissues was performed on a Leica Bond RX. All staining reagents were provided in the Opal 6-Plex Detection Kit (Akoya Bioscience, cat# NEL871001KT), which was used according to the manufacturer’s instructions for sequential staining of each antibody in the panel. Primary antibody information is detailed in **Supplementary Table 2**. Akoya Bioscience’s PhenoimagerHT platform and InForm® analysis software (v 2.8) was used for 20x whole slide scanning and spectral imaging of unmixed stains of 20x regions of interest. Tumor areas were manually annotated by the reference dermatopathologist (A.K.). Cell segmentations were generated using a Stardist model trained using a dataset of manual cell annotations from Vectra images. A multi-layer perceptron model was trained with a manually generated dataset of positive/negative annotations for a random selection of cells across all whole-slide images. This model was then used to classify cells as positive/negative for each marker. Each tumor mask in the Vectra dataset and each TMA core in the TMA dataset constituted a unique compartment for this analysis. We employed Ripley’s K function to assess the clustering dynamics of cells positive for each marker compared to all other markers within each compartment. SpatialTIME was used to calculate Ripley’s K for each compartment. Ripley’s bivariate *K* for a given radius *r* is defined as

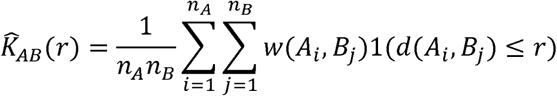

Where

*A_i_* is the *i*th cell positive for marker A

*B_i_* is the *j*th cell positive for marker B

*w*(*A_i,_ B_i_*) corrects for cells on the boundary of a compartment

*d*(*A_i,_ B_i_*) the Euclidean distance between cells *A_i_* and *B_i_*

For a random spatial distribution of cells 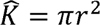 so the degree of clustering *D* can be calculated as 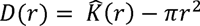. Negative values indicate a dispersion affect among cells between the two groups while positive values indicate a greater degree of clustering than expected from random chance.

### Peripheral blood flow cytometry

PBMCs were previously isolated from patient whole blood at designated experimental timepoints using Ficoll density gradient centrifugation. Five to 10 million PBMCs were resuspended in 1 ml 90%FBS/10%DMSO in cryovials, which were placed inside a Mr. Frosty unit and stored in the -80C freezer for 24-48 hours. Cryovials were then transferred to the vapor phase of a liquid nitrogen tank until study completion. In preparation for staining, PBMC cryovials were rapidly thawed in 37°C water bath until only a small piece of ice remained. One ml of warm RPMI was slowly dripped on top of the cells in the cryovial. The contents of the cryovial were pipetted slowly up and down and were then slowly transferred to a 15 ml conical tube containing 13 ml of warm RPMI. Tubes were then gently inverted 2-3 times and were centrifuged at 1500 rpm for 5 minutes to pellet PBMCs. Cells were resuspended in 1 ml of fresh RPMI and counted on a Nexcelom Cellometer. Approximately 500-600k viable patient PBMCs or healthy donor PMBCs were plated into individual wells of a 96-well round bottom plate for subsequent staining.

Plates were spun at 1500 rpm for 5 minutes to pellet cells. Supernatants were removed by quickly shucking the plate into a biohazard bag. Cells were washed in 100 ul of FACS buffer to remove traces of media. Plates were spun again at 1500 rpm for 5 minutes to pellet cells and the supernatants were shucked into a biohazard bag. Plate washing, spinning and shucking were performed between each staining step. Cells pellets were first stained with 50 ul of mixture of viability dye (Zombie NIR Fixable Viability Kit (Biolegend Inc 423105) and Human TruStain FcX™ (Biolegend Inc, 422302) for 15 min at room temperature in the dark. An antibody cocktail to surface markers was prepared in BD Horizon™ Brilliant Stain Buffer Plus (BD Biosciences, 566385) according to manufacturer’s instruction. Cells were stained in surface cocktail for 1 hour at 4C. Surface-stained cells were then fixed and permeabilized for 1 hour at 4C as per manufacturer instructions using the eBioscience™ Foxp3 / Transcription Factor Staining Buffer Set (ThermoFisher, 00-5523-00). A second antibody cocktail for intracellular markers (Ki67 and FOXP3) was prepared in 1x Perm Buffer from the aforementioned kit and cells were incubated overnight at 4C. The following morning, cells were washed in 100ul of 1x Perm Buffer. Cells were finally resuspended in 200 ul of FACS buffer and the data were acquired on the Cytek Aurora spectral flow cytometer. Data were analyzed using FlowJo v10.10.0 and Cytobank. FCS files were imported into Cytobank cloud-based platform. After performing normalization, debarcoding, and QC the processed data was analyzed via ViSNE tool for dimensional reduction and visualization, and FlowSOM was used for identifying clusters/metaclusters. The resulting t-SNE maps for various lead-in arms and response data were generated to show differences in immune subsets. Specifics for all antibodies and gating strategy used in this study can be found in the **Supplementary Table 3 and Fig. S9**.

### Single-cell RNAseq analyses

*CD8+ T cell signatures.* We sought to assess whether any gene signatures present in CD8+ T cells were associated with outcome following treatment. To determine this, we bioinformatically isolated CD8+ T cells from PBMC from our previously described datasets.^10^ From these CD8+ T cells, we identified clonally expanded CD8+ T cells as those that have at least 3 TCR beta CDR3 region amino acid sequences that were identical within a patient. We then performed gene set enrichment analyses using previously described genes sets^10^ as implemented by AddModuleScore in Seurat. When performed a linear mixed effects analysis as previously described^10^ in clonally expanded CD8+ T cells from blood to determine if any signatures across lead-in treatment groups were associated with pathological response.

*Ligand receptor analyses.* We used Celltalker^28^ to assess ligand receptor interactions in tumor infiltrating immune populations from our previously described single-cell RNAseq data from this cohort.^10^ Briefly, Celltalker uses known lists of ligands and receptors to look for interactions across cell types using single-cell RNAseq data. To determine the significance of interactions, Celltalker compares the joint mean expression of ligand and cognate receptors and compares to the joint mean of scrambled (i.e. not interacting) ligands and receptors. We used a previously described list of ligands/receptors for this analysis.^29^ Circos plots were used to visualize interactions across cell types using the circlize R package.

## List of Supplementary Materials

Fig. S1 Clinical trial schema Fig. S2 Clinical outcomes

Fig. S3 Pathologic response assessment using H&E

Fig. S4 Pathologic response parameters among 3 lead-in arms

Fig. S5 Lead-in therapy induced immune changes in tumor microenvironment Fig. S6 CD8^+^ T cell and FOXP3^+^ T cell interaction analysis

Fig. S7 Lead-in therapy induced CD4^+^ T cell, CD20^+^ B cell, CD68^+^ macrophage immune clustering and changes in tumor microenvironment

Fig. S8 CD33dim classical monocytes demonstrate a unique transcriptional signature. Fig. S9 Gating strategy of basic immune subpopulations.

Table S1 Treatment-related adverse number of events (percent) that occurred in ≥10% of patients and according to lead-in relatlimab (Rela), nivolumab (Nivo), and combination (Combo) therapy.

Table S2 Multiplex staining antibody panel. Table S3 Flow cytometry antibodies.

Data file S1. Adverse events among all patients.

## Author Contributions

Conceptualization: JMK, DAAV, TCB, LK, RCM, AR

Methodology: JMK, DAAV, TCB, LK, SW, HW, CS, ER, CD, RB

Investigation: JMK, LK, YGN, DD, JJL, SW, HW, RB

Visualization, image analyses: MJ, SK, AK

Funding acquisition: JMK, DAAV, TCB

Project administration: JMK

Supervision: JMK, DAAV, TCB

Writing – original draft: LK, JMK

Writing – review & editing: All authors

## Data Availability

All data produced in the present study are available upon reasonable request to the authors

## Acknowledgments

The authors would like to thank the Vignali lab (Vignali-lab.com; @Vignali_Lab), Bruno lab, Cillo lab for discussions and critically reading the manuscript. We would like to acknowledge all patients and their families for participating in this study.

## Funding

NIH-NCI (P50 SPORE CA254865 to JMK, DAAV, TCB)

Bristol-Myers Squibb (CA224-070 to JMK, TCB, and DAAV) We also thank Bristol-Myers Squibb for providing drug for this study.

University of Pittsburgh Center for Research Computing, RRID:SCR_022735, through the resources provided. Specifically, this work used the HTC cluster, which is supported by NIH award number S10OD028483. Bioinformatics analysis in the project described was performed by Cancer Bioinformatics Services (CBS), supported in part by NCI through the UPMC Hillman Cancer Center CCSG award (P30CA047904). This project used the Hillman Cancer Center Translational Pathology Imaging Laboratory that is supported in part by award P30CA047904 and the NSABP Foundation.

## Data and materials availability

All data is available in the main text or the supplementary materials.

