## Supplementary figures and tables for "Lead-in therapy targeting PD1 and/or LAG3 imposes distinct immune phenotypes in first-line treatment of metastatic melanoma"

**Supplementary tables for**

### Lead-in therapy targeting PD1 and/or LAG3 distinguishes differential impacts upon the immune response in first-line treatment of metastatic melanoma

Lilit Karapetyan[1.9], Anthony R. Cillo[2,3,4], Shuaichao Wang[5], Arivarasan Karunamurthy[6], Ryan C. Massa[1,10], Anjali Rohatgi[1,11], Christopher Deitrick[7], Yana G. Najjar[1,3], Diwakar Davar[1,3], Jason J. Luke[1,3], Cindy Sander[1], Sheryl R. Kunning[2,3], Amy Rose[1], Elizabeth Rush[1], Marion Joy[8], Riyue Bao[1,7], Hong Wang[5], Tullia C. Bruno[2,3,4,13], Dario A.A. Vignali[2,3,4,13], and John M. Kirkwood[1,3,13, 14]

**Affiliations**

[1]Department of Medicine, Division of Hematology/Oncology, University of Pittsburgh School of Medicine, Pittsburgh, PA, USA

[2]Department of Immunology, University of Pittsburgh, Pittsburgh, PA, USA

[3]Tumor Microenvironment Center, UPMC Hillman Cancer Center, University of Pittsburgh, Pittsburgh, PA, USA

[4]Cancer Immunology and Immunotherapy Program, UPMC Hillman Cancer Center, Pittsburgh, PA, USA

[5]UPMC Hillman Cancer Center Biostatistics Facility, Pittsburgh, PA, USA

[6]Department of Pathology, University of Pittsburgh School of Medicine, Pittsburgh, PA, USA

[7]UPMC Hillman Cancer Center Bioinformatics services, Pittsburgh, PA, USA

[8]Translational pathology imaging laboratory, UPMC Hillman Cancer Center, Pittsburgh, PA, USA

[9]H. Lee Moffitt Cancer Center & Research Institute, Tampa, FL, USA

[10]Abramson Cancer Center, Perelman School of Medicine, University of Pennsylvania, PA, USA

[11]Division of Oncology, Department of Medicine, Washington University School of Medicine, St Louis, MO, United States; Alvin J. Siteman Cancer Center, St Louis, MO, USA

[12]Center for Systems Immunology, Department of Immunology, University of Pittsburgh, Pittsburgh, PA USA

[13]Co-senior authors

[14]Lead contact

**Supplementary table 1.** Treatment-related adverse number of events (percent) that occurred in ≥10% of patients and according to lead-in relatlimab (Rela), nivolumab (Nivo), and combination (Combo) therapy.

| Adverse Event | Rela, N=14 | | Nivo, N=15 | | Combo, N=14 | | All, N=43 | |
|---|---|---|---|---|---|---|---|---|
| | *Any grade* | *Grade 3,4,5* | *Any grade* | *Grade 3,4,5* | *Any grade* | *Grade 3,4,5* | *Any grade* | *Grade 3,4,5* |
| Any adverse event | 14 (100) | 13 (92.9) | 15 (100) | 12 (80) | 14 (100) | 9 (64.3) | 43 (100) | 34 (79.1) |
| Treatment-related adverse event | 10 (71.4) | 6 (42.9) | 13 (86.7) | 7 (46.7) | 13 (92.9) | 2 (14.3) | 36 (83.7) | 15 (34.9) |
| Treatment-related adverse event in 10% of patients | | | | | | | | |
| Fatigue | 7 (50) | 0 (0) | 7 (46.7) | 1 (6.7) | 3 (21.4) | 0 (0) | 17 (39.5) | 1 (2.3) |
| Hyponatremia | 6 (42.9) | 1 (7.1) | 8 (53.3) | 3 (20) | 3 (21.4) | 0 (0) | 17 (39.5) | 4 (9.3) |
| Serum amylase increased | 6 (42.9) | 3 (21.4) | 3 (20) | 0 (0) | 6 (42.9) | 0 (0) | 15 (34.9) | 3 (7) |
| Aspartate aminotransferase increased | 5 (35.7) | 0 (0) | 4 (26.7) | 1 (6.7) | 3 (21.4) | 0 (0) | 12 (27.9) | 1 (2.3) |
| Alanine aminotransferase increased | 5 (35.7) | 0 (0) | 4 (26.7) | 1 (6.7) | 2 (14.3) | 0 (0) | 11 (25.6) | 1 (2.3) |
| Hypothyroidism | 3 (21.4) | 0 (0) | 3 (20) | 0 (0) | 3 (21.4) | 0 (0) | 9 (20.9) | 0 (0) |
| Hypophosphatemia | 1 (7.1) | 0 (0) | 4 (26.7) | 0 (0) | 4 (28.6) | 0 (0) | 9 (20.9) | 0 (0) |
| Rash maculo-papular | 2 (14.3) | 0 (0) | 3 (20) | 0 (0) | 4 (28.6) | 0 (0) | 9 (20.9) | 0 (0) |
| Nausea | 3 (21.4) | 0 (0) | 3 (20) | 0 (0) | 2 (14.3) | 0 (0) | 8 (18.6) | 0 (0) |
| Lipase increased | 3 (21.4) | 2 (14.3) | 3 (20) | 1 (6.7) | 2 (14.3) | 1 (7.1) | 8 (18.6) | 4 (9.3) |
| Thyroid stimulating hormone increased | 3 (21.4) | 0 (0) | 3 (20) | 0 (0) | 2 (14.3) | 0 (0) | 8 (18.6) | 0 (0) |
| Adrenal insufficiency | 3 (21.4) | 1 (7.1) | 2 (13.3) | 0 (0) | 2 (14.3) | 0 (0) | 7 (16.3) | 1 (2.3) |
| Alkaline phosphatase increased | 1 (7.1) | 0 (0) | 3 (20) | 1 (6.7) | 3 (21.4) | 0 (0) | 7 (16.3) | 1 (2.3) |
| Pruritus | 1 (7.1) | 1 (7.1) | 2 (13.3) | 0 (0) | 3 (21.4) | 0 (0) | 6 (14) | 1 (2.3) |
| Neutrophil count decreased | 0 (0) | 0 (0) | 4 (26.7) | 0 (0) | 1 (7.1) | 0 (0) | 5 (11.6) | 0 (0) |

Note: Colitis: 3 patients experienced colitis of any grade; Encephalitis: 1 patient had encephalitis; Myocarditis: 1 patient had myocarditis.

#### Supplementary Table 2. Multiplex staining antibody panel

| Manufacturer | product# | Target | clone | AR | Block | 2° HRP | Opal |
|---|---|---|---|---|---|---|---|
| Biocare | API3209AA | CD4 | EP204 | 9 | Akoya | Akoya | 690 |
| Cell Signaling | 76437S | CD68 | D4B96 | 6 | Akoya | Akoya | 520 |
| Cell Signaling | 12653S | FoxP3 | D608R | 6 | Akoya | Akoya | 570 |
| Leica | NC-L-CD20-L26 | CD20 | L26 | 6 | Akoya | Akoya | 620 |
| BioCare | ACI3160A | CD8 | C8/144B | 6 | Akoya | **Leica PowerVision Poly-HRP Anti-Mouse IgG** | 480 |
| Abcam | ab212843 | SOX10 | SOX10/991 | 6 | Akoya | Akoya | 780 |

**Supplementary Table 3. Flow cytometry antibodies**

| | Fluorophore | Antigen | Dilution Factor | Vendor | Cat# | Clone |
|---|---|---|---|---|---|---|
| Surface | APC-Fire810 | CD19 | 50 | BL | 302272 | HIB19 |
| | BV510 | CD1c | 50 | BL | 331534 | L-161 |
| | BV605 | CD62L | 100 | BD | 562719 | DREG-56 |
| | BV650 | CD163 | 100 | BD | 563888 | GHI/61 |
| | BV785 | CD141 | 100 | BL | 344116 | M80 |
| | APC | CD138 | 100 | BL | 356506 | M115 |
| | Spark Blue 550 | CD3 | 100 | BL | 344852 | SK7 |
| | PE-Cy7 | CCR7 | 150 | BL | 353226 | G043H7 |
| | BV480 | CD38 | 150 | BD | 566137 | HIT2 |
| | BV711 | CD25 | 150 | BL | 302636 | BC96 |
| | PerCP-Cy5.5 | CD11b | 150 | BL | 301328 | ICRF44 |
| | PE-Cy5 | CD27 | 150 | TF | 15-0279-42 | 323 |
| | Alexa700 | CD11c | 150 | BL | 301648 | 3.9 |
| | BV750 | CD20 | 150 | BD | 747062 | 2H7 |
| | PE-Dazz | CD16 | 150 | BL | 302054 | 3G8 |
| | BUV615 | CD103 | 300 | BD | 751258 | Ber-ACT8 |
| | BUV737 | CD14 | 300 | BD | 612763 | M5E2 |
| | APC-Cy7 | CD66B | 300 | BL | 305126 | G10F5 |
| | BUV 661 | CD56 | 400 | BD | 750478 | NCAM16.2 |
| | BUV496 | CD8 | 400 | BD | 612942 | RPA-T8 |
| | BV570 | CD45RA | 600 | BL | 304132 | H1100 |
| | BUV563 | CD4 | 600 | BD | 741353 | RPA-T4 |
| | BUV395 | CD45 | 600 | BD | 563792 | HI30 |
| | AF488 | HLADR | 800 | BL | 307620 | L243 |
| | PE | CD33 | 1000 | BL | 366608 | P67.6 |
| ICS | BV421 | Ki67 | 200 | BL | 350506 | Ki-67 |
| | eFluor450 | FoxP3 | 100 | TF | 48-4776-42 | PCH101 |

**Supplementary figures for**

### Lead-in therapy targeting PD1 and/or LAG3 imposes distinct immune phenotypes in first-line treatment of metastatic melanoma

Lilit Karapetyan[1,9], Anthony R. Cillo[2,3,4], Shuaichao Wang[5], Arivarasan Karunamurthy[6], Ryan C. Massa[1,10], Anjali Rohatgi[1,11], Christopher Deitrick[7], Yana G. Najjar[1,3], Diwakar Davar[1,3], Jason J. Luke[1,3], Cindy Sander[1], Sheryl R. Kunning[2,3], Amy Rose[1], Sarah Bradley[2,3,4], Elizabeth Rush[1], Marion Joy[8], Riyue Bao[1,7], Hong Wang[5], Dario A.A. Vignali[2,3,4,13], Tullia C. Bruno[2,3,4,13], and John M. Kirkwood[1,3,13, 14]

**Affiliations**

[1]Department of Medicine, Division of Hematology/Oncology, University of Pittsburgh School of Medicine, Pittsburgh, PA, USA

[2]Department of Immunology, University of Pittsburgh, Pittsburgh, PA, USA

[3]Tumor Microenvironment Center, UPMC Hillman Cancer Center, University of Pittsburgh, Pittsburgh, PA, USA

[4]Cancer Immunology and Immunotherapy Program, UPMC Hillman Cancer Center, Pittsburgh, PA, USA

[5]UPMC Hillman Cancer Center Biostatistics Facility, Pittsburgh, PA, USA

[6]Department of Pathology, University of Pittsburgh School of Medicine, Pittsburgh, PA, USA

[7]UPMC Hillman Cancer Center Bioinformatics services, Pittsburgh, PA, USA

[8]Translational pathology imaging laboratory, UPMC Hillman Cancer Center, Pittsburgh, PA, USA

[9]H. Lee Moffitt Cancer Center & Research Institute, Tampa, FL, USA

[10]Abramson Cancer Center, Perelman School of Medicine, University of Pennsylvania, PA, USA

[11]Division of Oncology, Department of Medicine, Washington University School of Medicine, St Louis, MO, United States; Alvin J. Siteman Cancer Center, St Louis, MO, USA

[12]Center for Systems Immunology, Department of Immunology, University of Pittsburgh, Pittsburgh, PA USA

[13]Co-senior authors

[14]Lead contact

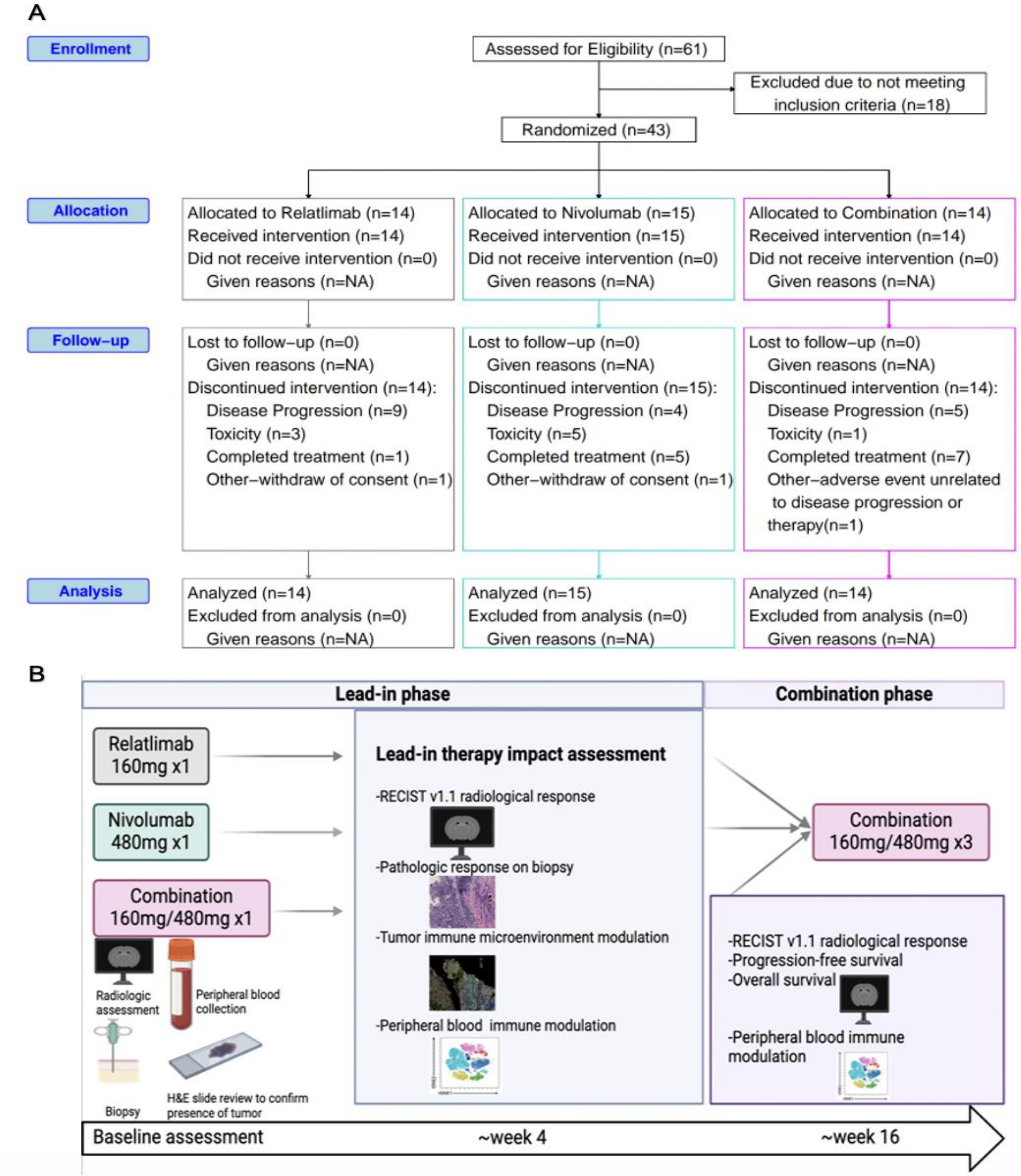


**Supplementary figure 1: Clinical trial schema. (A)** CONSORT diagram depicting patient screening and disposition. (**B)** Clinical trial schema depicting assessment timepoints for radiological response, pathologic response, tumor immune microenvironment and peripheral blood immunophenotyping changes in lead-in and combination-phases.

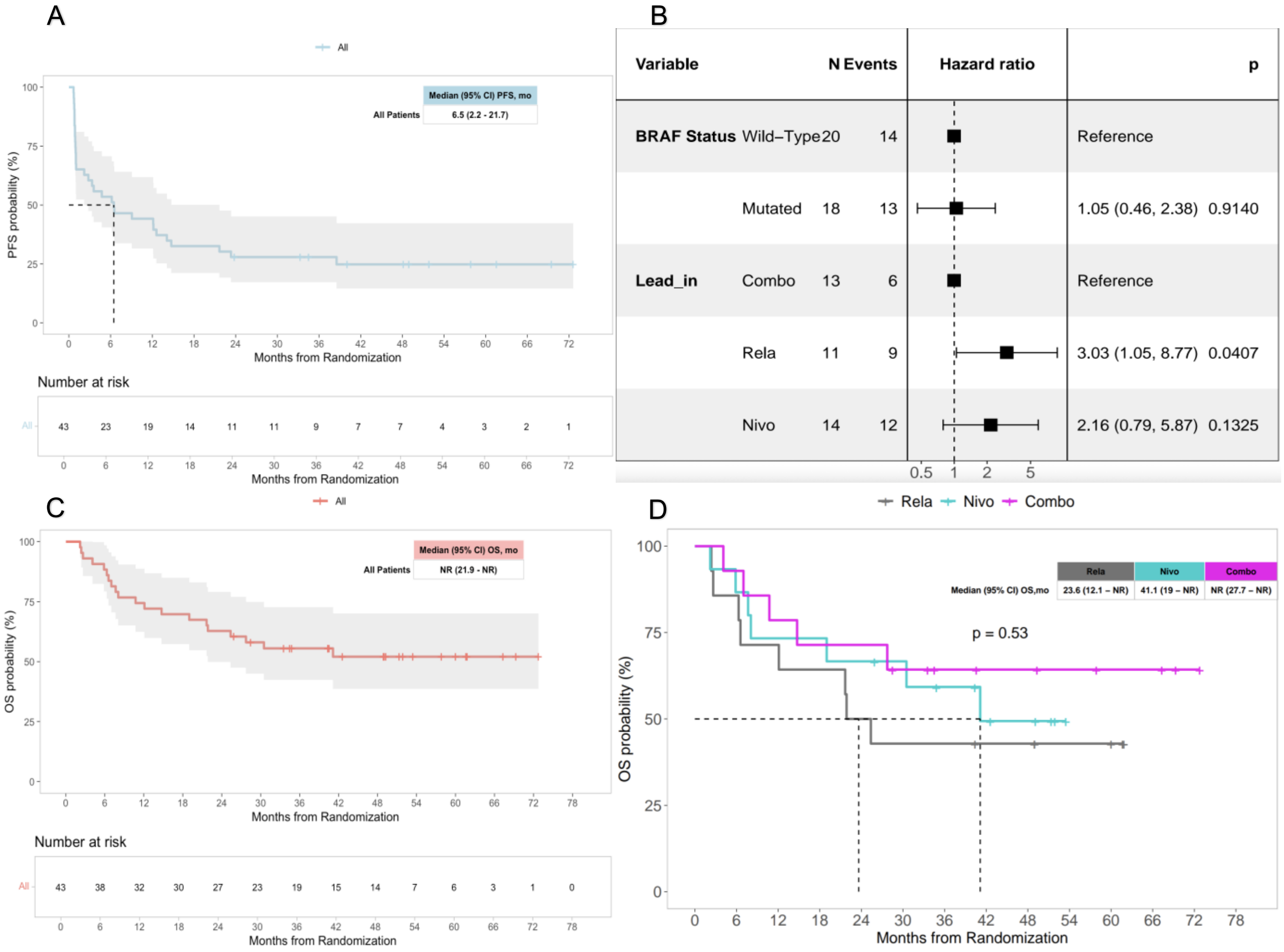
A
All
Median (95% CI) PFS, mo
All Patients
6.5 (2.2 - 21.7)
PFS probability (%)
Months from Randomization
Number at risk
43 23 19 14 11 11 9 7 7 4 3 2 1
B
Variable
N
Events
Hazard ratio
p
BRAF Status
Wild–Type
20
14
Reference
Mutated
18
13
1.05 (0.46, 2.38)
0.9140
Lead_in
Combo
13
6
Reference
Rela
11
9
3.03 (1.05, 8.77)
0.0407
Nivo
14
12
2.16 (0.79, 5.87)
0.1325
C
All
Median (95% CI) OS, mo
All Patients
NR (21.9 - NR)
OS probability (%)
Months from Randomization
Number at risk
43 38 32 30 27 23 19 15 14 7 6 3 1 0
D
Rela
Nivo
Combo
Median (95% CI) OS,mo
23.6 (12.1 – NR)
41.1 (19 – NR)
NR (27.7 – NR)
p = 0.53
OS probability (%)
Months from Randomization

**Supplementary figure 2: Clinical outcomes (A)** Progression-free survival (PFS) among all patients. (**B)** Forest Plot for Cox proportional hazards model showing the impact of lead-in therapy after adjusting for *BRAFv600* mutation status. **(C)** Overall survival (OS) among all patients. (**D)** OS according to lead-in arm.

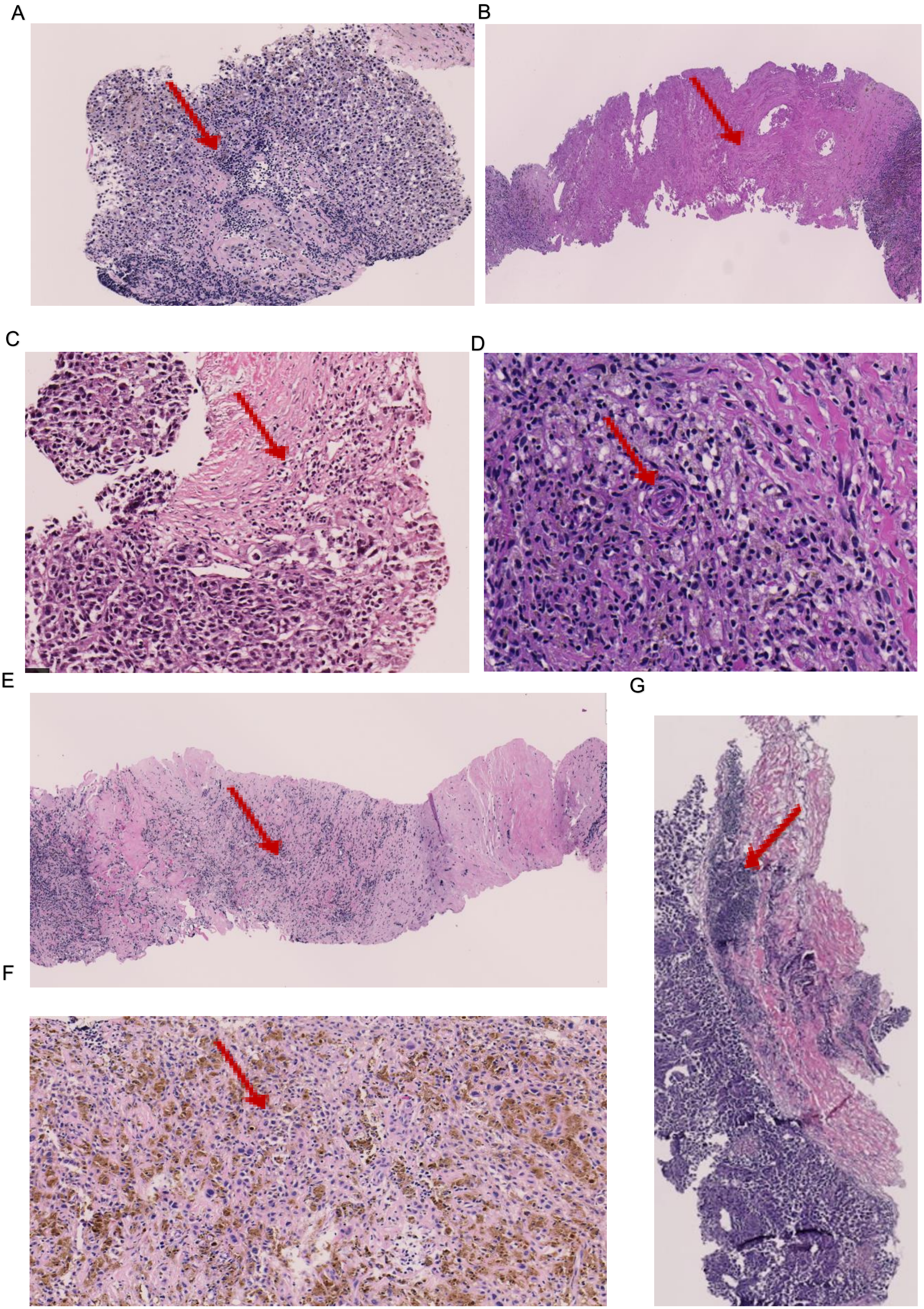
A
B
C
D
E
G
F

**Supplementary figure 3: Pathologic response assessment using H&E. (A)** tumor-infiltrating lymphocytes. (**B)** absence of viable tumor. (**C)** plasma cells. (**D)** neovascularization. (**E)** fibrosis. (**F)** viable tumor with melanosis. (**G)** lymphoid aggregates.

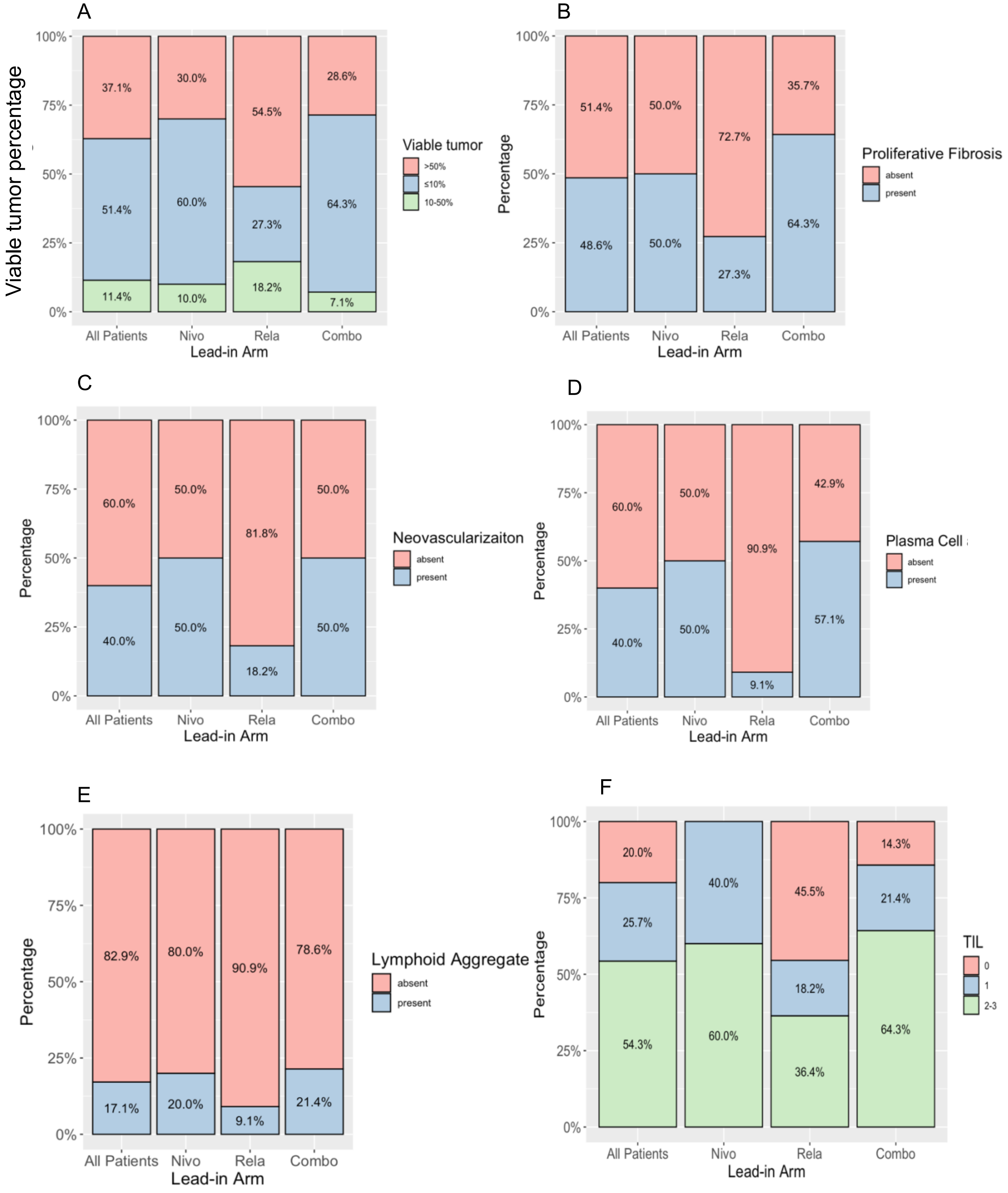

A
Viable tumor percentage
100%
75%
50%
25%
0%
37.1%
30.0%
54.5%
28.6%
51.4%
60.0%
27.3%
64.3%
11.4%
10.0%
18.2%
7.1%
Viable tumor
>50%
≤10%
10-50%
All Patients
Nivo
Rela
Combo
Lead-in Arm
B
Percentage
51.4%
50.0%
72.7%
35.7%
48.6%
50.0%
27.3%
64.3%
Proliferative Fibrosis
absent
present
All Patients
Nivo
Rela
Combo
Lead-in Arm
C
Percentage
60.0%
50.0%
81.8%
50.0%
40.0%
50.0%
18.2%
50.0%
Neovascularizaiton
absent
present
All Patients
Nivo
Rela
Combo
Lead-in Arm
D
Percentage
60.0%
50.0%
90.9%
42.9%
40.0%
50.0%
9.1%
57.1%
Plasma Cell
absent
present
All Patients
Nivo
Rela
Combo
Lead-in Arm
E
Percentage
82.9%
80.0%
90.9%
78.6%
17.1%
20.0%
9.1%
21.4%
Lymphoid Aggregate
absent
present
All Patients
Nivo
Rela
Combo
Lead-in Arm
F
Percentage
20.0%
25.7%
54.3%
40.0%
60.0%
45.5%
18.2%
36.4%
14.3%
21.4%
64.3%
TIL
0
1
2-3
All Patients
Nivo
Rela
Combo
Lead-in Arm

**Supplementary figure 4: Pathologic response parameters among 3 lead-in arms.** The percentages are depicted among all patients and nivolumab (nivo), relatlimab (rela), and combination (combo) arms, respectively. **(A)** residual viable tumor. (**B)** proliferative fibrosis (present/absent). (**C)** neovascularization (present/absent). (**D)** plasma cells (present/absent). (**E)** lymphoid aggregate (present/absent). (**F)** tumor-infiltrating lymphocytes (0 vs 1 vs 2-3).

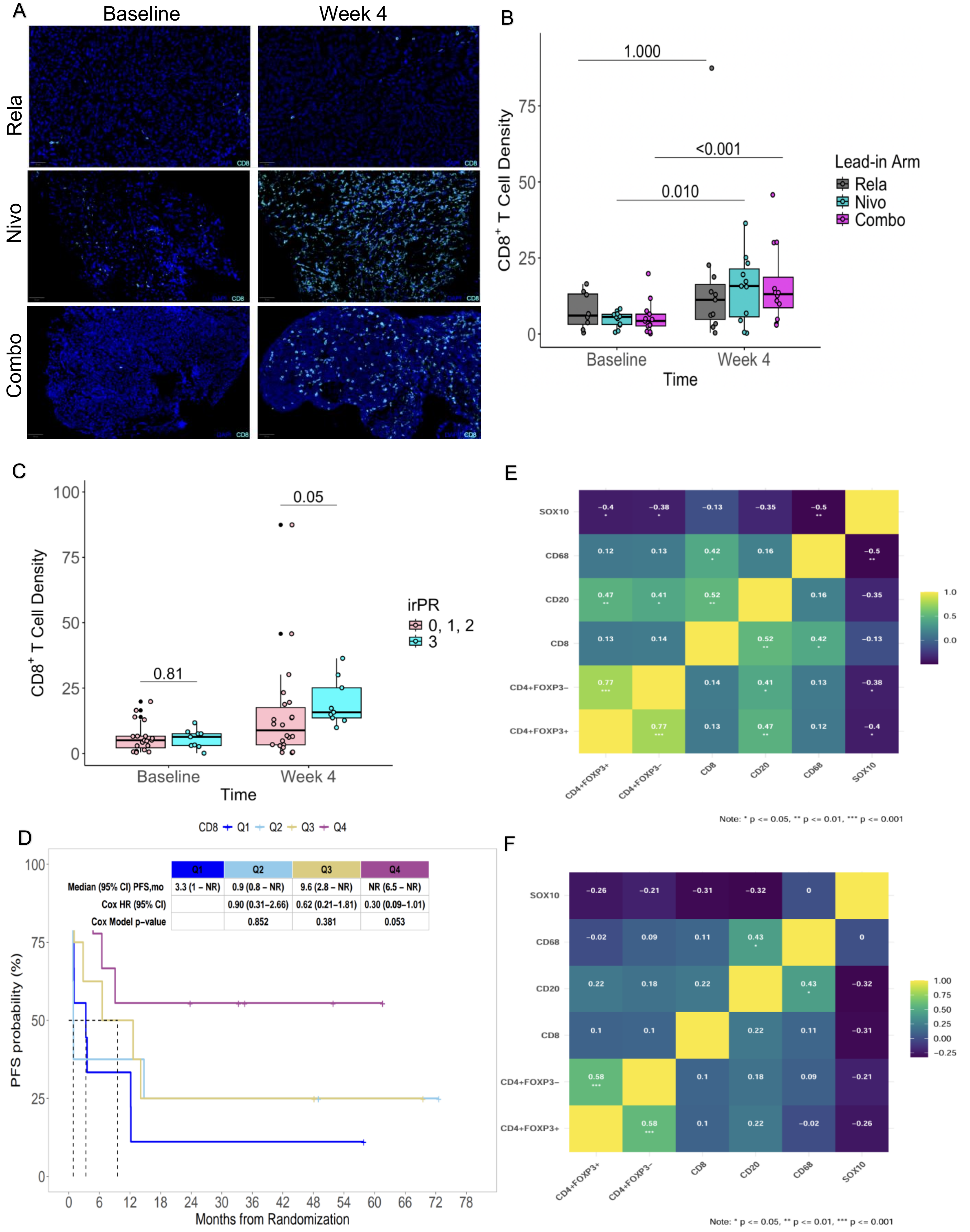
A
Baseline
Week 4
Rela
Nivo
Combo
B
1.000
<0.001
0.010
CD8+ T Cell Density
Baseline
Week 4
Time
Lead-in Arm
Rela
Nivo
Combo
C
0.05
0.81
CD8+ T Cell Density
irPR
0, 1, 2
3
Baseline
Week 4
Time
D
CD8
Q1
Q2
Q3
Q4
Median (95% CI) PFS,mo
3.3 (1 – NR)
0.9 (0.8 – NR)
9.6 (2.8 – NR)
NR (6.5 – NR)
Cox HR (95% CI)
0.90 (0.31–2.66)
0.62 (0.21–1.81)
0.30 (0.09–1.01)
Cox Model p-value
0.852
0.381
0.053
PFS probability (%)
Months from Randomization
E
SOX10
CD68
CD20
CD8
CD4+FOXP3–
CD4+FOXP3+
Note: * p <= 0.05, ** p <= 0.01, *** p <= 0.001
F
SOX10
CD68
CD20
CD8
CD4+FOXP3–
CD4+FOXP3+
Note: * p <= 0.05, ** p <= 0.01, *** p <= 0.001

**Supplementary figure 5: Lead-in therapy induced immune changes in tumor microenvironment. (A)** Combination (combo) and nivolumab (nivo) but not relatlimab (rela) lead-in therapies led to significant increase in $CD8^+$ T cell density. (**B)** The comparison was performed between baseline and week 4 data. p values were calculated using Wilcoxon signed-rank test. (**C)** $CD8^+$ T cell density at baseline and week 4 in association with immune related pathologic response (irPR). Box plot demonstrates significant increase in week 4 $CD8^+$ T cell density among irPR=3 (major pathologic response on biopsy) vs irPR=0,1,2. (**D)** PFS according to $CD8^+$ T cell density at week 4. (**E)** Correlation plot of cell densities at baseline. (**F)** week 4 samples. Correlation coefficient and p values are depicted in the figure.

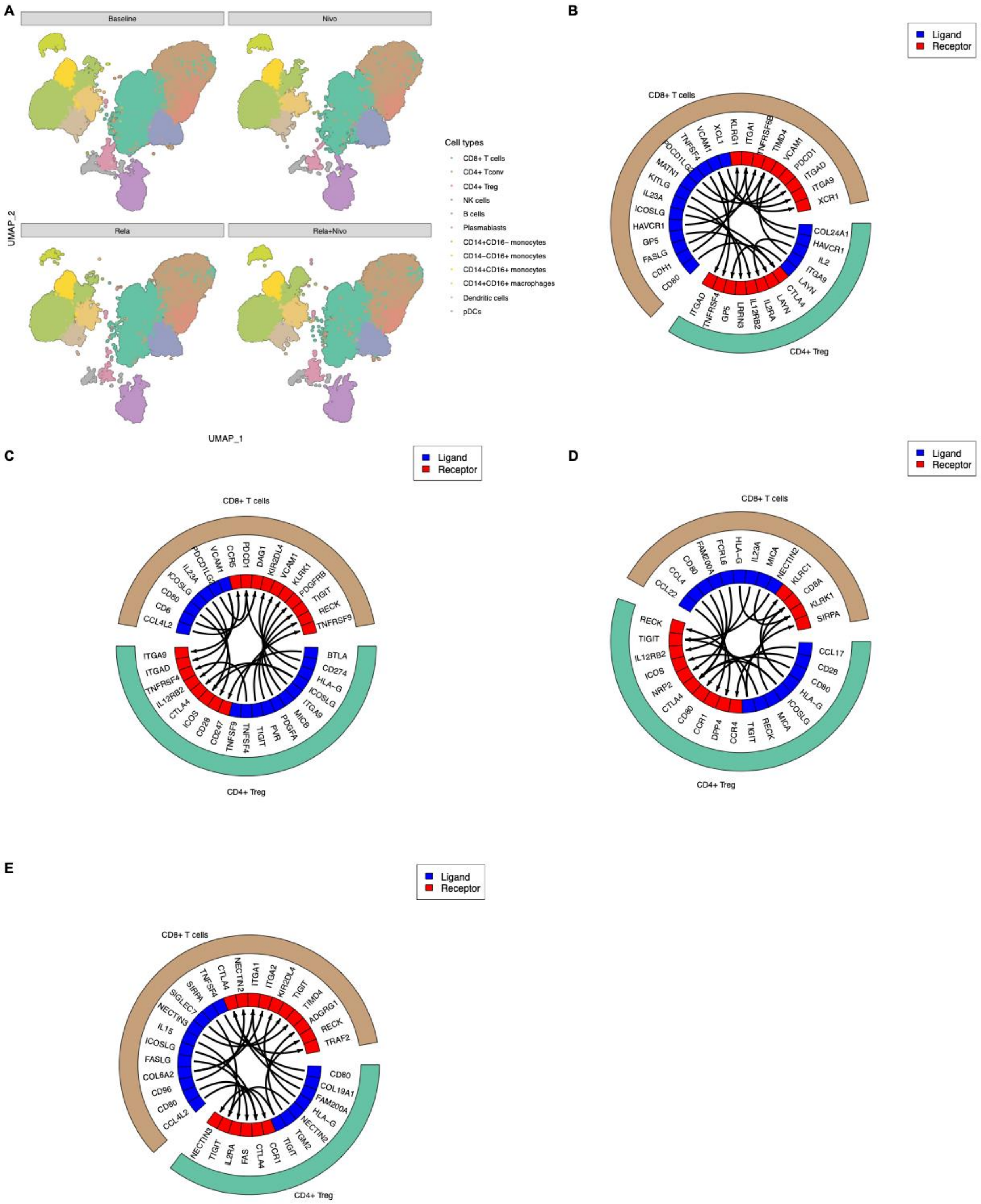
A
Baseline
Nivo
Rela
Rela+Nivo
UMAP_2
UMAP_1
Cell types
CD8+ T cells
CD4+ Tconv
CD4+ Treg
NK cells
B cells
Plasmablasts
CD14+CD16− monocytes
CD14−CD16+ monocytes
CD14+CD16+ monocytes
CD14+CD16+ macrophages
Dendritic cells
pDCs
B
Ligand
Receptor
CD8+ T cells
CD4+ Treg
C
Ligand
Receptor
CD8+ T cells
CD4+ Treg
D
Ligand
Receptor
CD8+ T cells
CD4+ Treg
E
Ligand
Receptor
CD8+ T cells
CD4+ Treg

**Supplementary figure 6: CD8$^+$ T cell and FOXP3$^+$ T cell interaction analysis.** (**A)** UMAP plot illustrating immune cell subsets identified within tumor immune microenvironment at baseline and week 4 across 3 lead-in arms. (**B)** Circos plot for top ligand-receptor interactions at baseline. (**C)** At week 4 in relatlimab (Rela) lead-in arm. (**D)** Nivolumab (Nivo) lead-in arm. (**E)** Combination (Combo) lead-in arm.

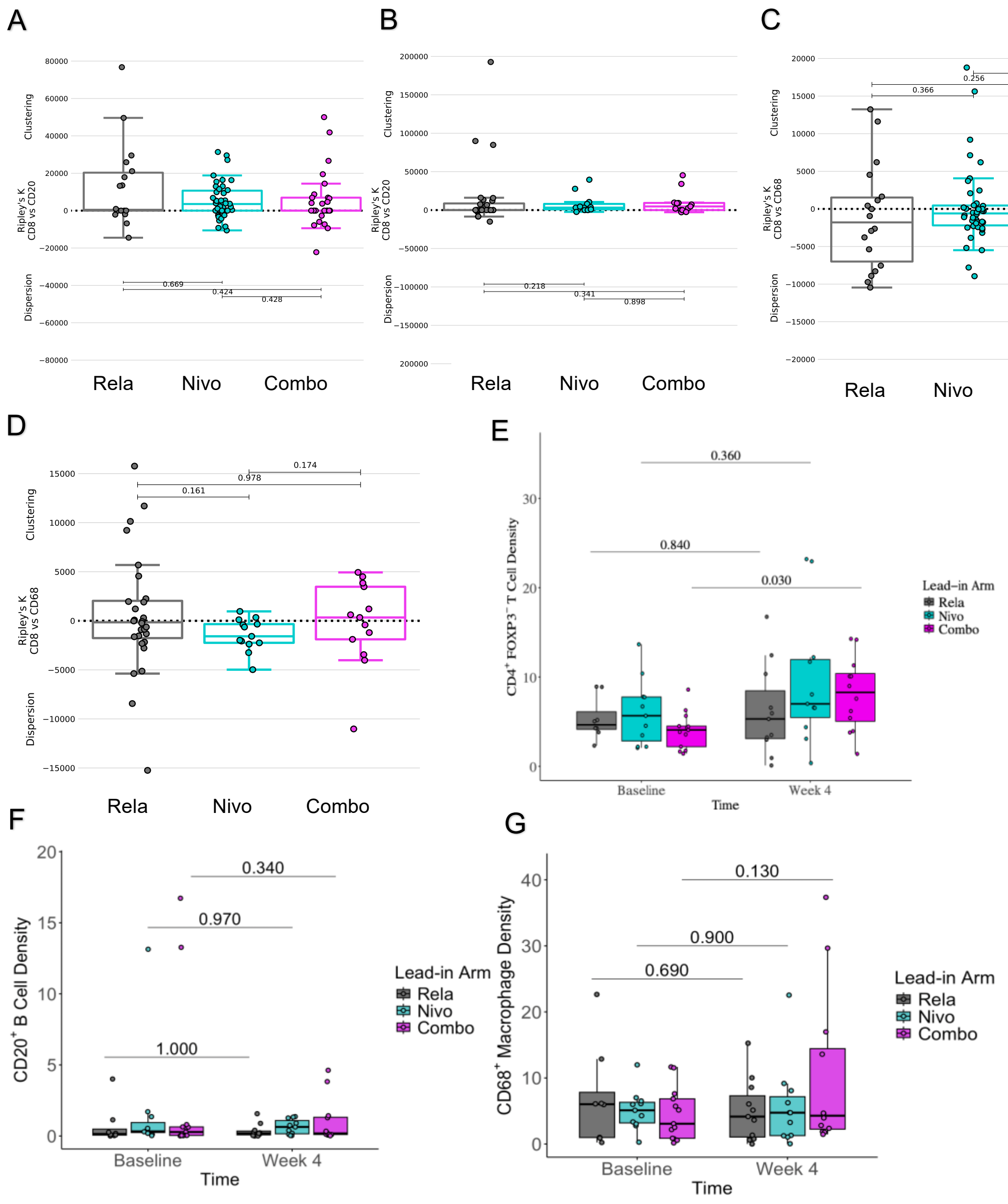
A
Clustering
Ripley's K CD8 vs CD20
Dispersion
0.669
0.424
0.428
Rela
Nivo
Combo
B
Clustering
Ripley's K CD8 vs CD20
Dispersion
0.218
0.341
0.898
Rela
Nivo
Combo
C
Clustering
Ripley's K CD8 vs CD68
Dispersion
0.363
0.256
0.366
Rela
Nivo
Combo
D
Clustering
Ripley's K CD8 vs CD68
Dispersion
0.174
0.978
0.161
Rela
Nivo
Combo
E
CD4+ FOXP3− T Cell Density
0.360
0.840
0.030
Lead-in Arm
Rela
Nivo
Combo
Baseline
Week 4
Time
F
CD20+ B Cell Density
0.340
0.970
1.000
Lead-in Arm
Rela
Nivo
Combo
Baseline
Week 4
Time
G
CD68+ Macrophage Density
0.130
0.900
0.690
Lead-in Arm
Rela
Nivo
Combo
Baseline
Week 4
Time

**Supplementary figure 7: Lead-in therapy induced $CD4^+$ T cell, $CD20^+$ B cell, $CD68^+$ macrophage immune clustering and changes in tumor microenvironment. (A)** Box plot indicates baseline colocalization patterns of $CD8^+$ T cells and $CD20^+$ B cells. Ripley's K function is illustrated on y axis. Higher numbers indicate clustering and lower numbers indicate dispersion of these two-cell types. The comparison of baseline samples among 3 lead-in arms reveals no significant differences in colocalization patterns. P values were calculated using a Wilcoxon rank-sum test. (**B)** Box plot indicates week 4 colocalization patterns of $CD8^+$ T cells and $CD20^+$ B cells. The comparison of week 4 samples among 3 lead-in arms reveals no significant differences in colocalization patterns. P values were calculated using a Wilcoxon rank-sum test. (**C)** Box plot indicates baseline colocalization patterns of $CD8^+$ T cells and $CD68^+$ macrophages. Ripley's K function is illustrated on y axis. Higher numbers indicate clustering and lower numbers indicate dispersion of these two-cell types. The comparison of baseline samples among 3 lead-in arms reveals no significant differences in colocalization patterns. P values were calculated using a Wilcoxon rank-sum test. (**D)** Box plot indicates week 4 colocalization patterns of $CD8^+$ T cells and $CD68^+$ macrophages. Ripley's K function is illustrated on y axis. Higher numbers indicate clustering and lower numbers indicate dispersion of these two-cell types. The comparison of week 4 samples among 3 lead-in arms reveals no significant differences in colocalization patterns. P values were calculated using a Wilcoxon rank-sum test. (**E)** Combo but not nivo or rela lead-in therapy led to significant increase in $CD4^+FOXP3^-$ T cells. p values were calculated using Wilcoxon signed-rank test. (**F)** No significant changes were observed on $CD20^+$ B cell density among 3 lead-in arms. (**G)** No significant changes were observed on $CD68^+$ macrophage density among 3 lead-in arms.

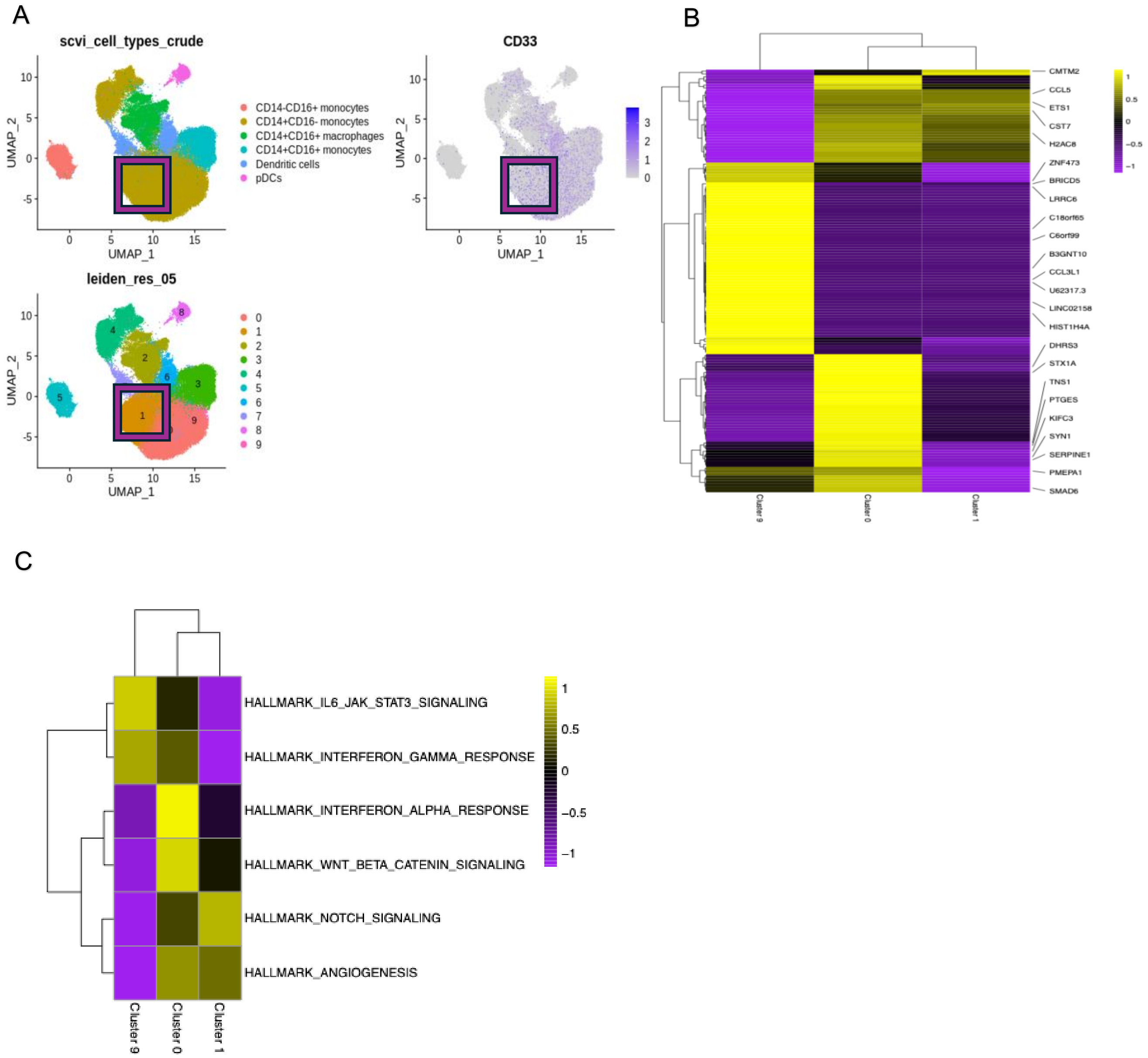
A
scvi_cell_types_crude
CD14-CD16+ monocytes
CD14+CD16- monocytes
CD14+CD16+ macrophages
CD14+CD16+ monocytes
Dendritic cells
pDCs
UMAP_1
UMAP_2
CD33
leiden_res_05
B
CMTM2
CCL5
ETS1
CST7
H2AC8
ZNF473
BRICD5
LRRC6
C18orf65
C6orf99
B3GNT10
CCL3L1
U62317.3
LINC02158
HIST1H4A
DHRS3
STX1A
TNS1
PTGES
KIFC3
SYN1
SERPINE1
PMEPA1
SMAD6
Cluster 9
Cluster 0
Cluster 1
C
HALLMARK_IL6_JAK_STAT3_SIGNALING
HALLMARK_INTERFERON_GAMMA_RESPONSE
HALLMARK_INTERFERON_ALPHA_RESPONSE
HALLMARK_WNT_BETA_CATENIN_SIGNALING
HALLMARK_NOTCH_SIGNALING
HALLMARK_ANGIOGENESIS
Cluster 9
Cluster 0
Cluster 1

**Supplementary figure 8: CD33dim classical monocytes demonstrate a unique transcriptional signature. (A)** Leiden clustering identified 9 distinct clusters (clusters 0 through 9) of myeloid cells in PBMC. Square highlights CD33 dim classical monocytes (cluster 1). (**B)** Heat map reveals differentially expressed genes across classical monocytes (clusters 0, 1, 9). (**C)** Gene set enrichment analysis across classical monocytes (clusters 0, 1, 9).

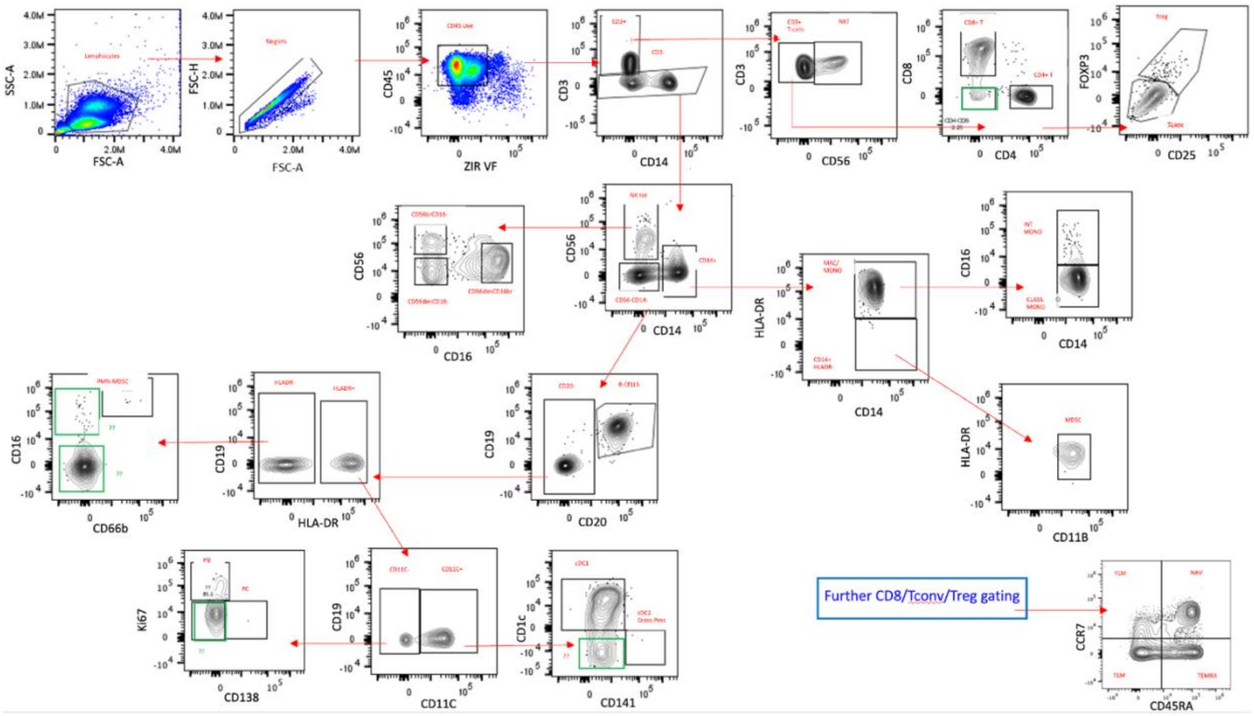

**Supplementary figure 9: Gating strategy of basic immune subpopulations.**
